# An App-Based Ecological Momentary Assessment of Undergraduate Mental Health During the COVID-19 pandemic in Canada (Smart Healthy Campus Version 2.0): Longitudinal Study

**DOI:** 10.1101/2023.03.22.23287598

**Authors:** Chris Brogly, Daniel J Lizotte, Marc Mitchell, Mark Speechley, Arlene MacDougall, Erin Huner, Kelly K Anderson, Michael A Bauer

**Affiliations:** Faculty of Information and Media Studies, Western University, London, ON, CA; Faculty of Health Sciences, Western University, London, ON, CA; Department of Computer Science, Western University, London, ON, CA; Department of Epidemiology and Biostatistics, Schulich School of Medicine and Dentistry, Western University, London, ON, CA; School of Kinesiology, Faculty of Health Sciences, Western University, London, ON, CA; Department of Psychiatry, Schulich School of Medicine and Dentistry, Western University, London, ON, CA; Ivey Business School, Western University, London, ON, CA

## Abstract

This paper presents results from the Smart Healthy Campus 2.0 study/smartphone app, developed and used to collect mental health-related lifestyle data from 86 Canadian undergraduates January – August 2021. This was a longitudinal repeat measures study conducted over 40 weeks. A 9-item mental health questionnaire was accessible once daily in the app. Two variants of this mental health questionnaire existed; the first was a weekly variant, available each Monday or until a participant responded during the week. The second was a daily variant available after the weekly variant. Mixed models were fit for responses to the two variants and 12 phone digital measures (e.g. GPS, step counts). A second round of models was fit based on backward elimination to determine associations between the 12 digital measures and the variants. 6518 digital measure samples and 1722 questionnaire responses were collected. The daily questionnaire had positive associations with floors walked, installed apps, and campus proximity, while having negative associations with uptime, and daily calendar events. Daily depression had a positive association with uptime. Daily resilience appeared to have a slight positive association with campus proximity. The weekly questionnaire variant had positive associations with device idling and installed apps, and negative associations with floors walked, calendar events, and campus proximity. Physical activity, weekly, had a negative association with uptime, and a positive association with calendar events and device idling. SHC 2.0, via phone digital measures, identified indicators of lifestyle that appeared to be associated with measures of mental health in undergraduates during COVID-19.

**Author Summary:** This paper analyzes the associations between digital measures from smartphones (such as GPS and step counts) and a broad mental health questionnaire (covering items like depression and anxiety). This data was collected from students at a relatively large, urban university in Canada during the COVID-19 pandemic. We conducted this study because smartphone-based studies observing aspects of mental health in students as they go about their daily lives are uncommon in Canada. Additionally, mental health concerns, such as depression and anxiety, can be common on university campuses, although it isn’t always clear what is associated with those concerns, especially when only looking at them with smartphone data. We were also interested in how an overview of student mental health would relate to digital measures coming from smartphones during the pandemic, as this information would be relevant to inform future pandemics. In general, these relationships might potentially inform ways to improve student mental health, or potentially predict aspects of it, based on data coming in from smartphones.

## Introduction

Mental health issues on university campuses are common during periods of traditional in-person study ^1^. Universities in Canada ^2^ have recognized that undergraduate students are faced with a number of stresses that challenge mental health. Undergraduate studies are demanding, and students often feel overwhelmed by their obligations ^1^. Additionally, the COVID-19 pandemic had an unprecedented effect on post-secondary students all over Canada, who experienced disruptive ^3–7^ changes to their studies in response to necessary public health measures regarding COVID-19. Canadian undergraduates have been surveyed regarding aspects of mental health using traditional sampling techniques such as questionnaires and interviews on several occasions ^8–10^. Another sampling technique particularly useful to this area is the Ecological Momentary Assessment (EMA) which allows for students to be sampled regarding aspects of their mental health as they go about their days; EMA studies are generally facilitated via mobile apps that run on Android and iOS. In addition to questionnaire responses, the apps can collect a wealth of data related to mental health from device hardware, such as GPS location, step counts, and indicators of life activity. For instance, the StudentLife study from Dartmouth College was one of the first in this area to assess student mental health using an app^11^.

We were interested in studying aspects of student mental health in general via mobile apps as, to the best of the author’s knowledge, little work has been done in this area at Canadian universities. We also had a particular interest in how mental health measures might be associated with various digital measures coming from smartphone hardware. As a result, we developed and launched the Smart Healthy Campus 2.0 app on Android and iOS. This ran on a research platform we called EMAX, which has been outlined in a previous paper ^12^. SHC 2.0 is the successor to a limited, pilot SHC 1.0 study, also detailed in a previous paper ^13^. The SHC 2.0 app asks a small number of mental health-related questions on a daily basis and collects data from phone hardware as questionnaires are submitted and also occasionally in the background. It was designed to be run during traditional on-campus study for an extended period, such as several months or even semesters. However, the SHC 2.0 study ended up being launched and run during a significant portion of the COVID-19 pandemic, as it was still relevant. We also launched a separate, strongly-pandemic focused study/app called Student Pandemic Experience (SPE) ^14^, which collected more data than the SHC 2.0 study, but was not suitable for use past the pandemic due to the specialized questionnaires used. For SHC 2.0, objectives of the study were to 1) address the lack of longitudinal data in Canada on overviews of student mental health and any potentially related measures of lifestyle, and 2) to identify associations between these self-reported overviews of mental health (from questionnaires) and lifestyle-related measures (from smartphone digital measures). In this study, we focused on providing a more complete analysis of any and all device digital measures that might relate to student mental health rather than focusing heavily on specific ones. Available smartphone digital measures (such as GPS, step counts) were still categorized into related groups, informed by previous work in mobile sensing^15^, including 1) Movement and physical activity, 2) Device usage, and 3) Social and life activity indicators. As this was exploratory observational research, we did not make specific hypotheses about how these various phone sensors might explain aspects of participant mental health – we were only interested in the prospect of whether any associations between the two might exist. We consequently decided not to elaborate on the meaning or function of any associations.

**Figure 1.**
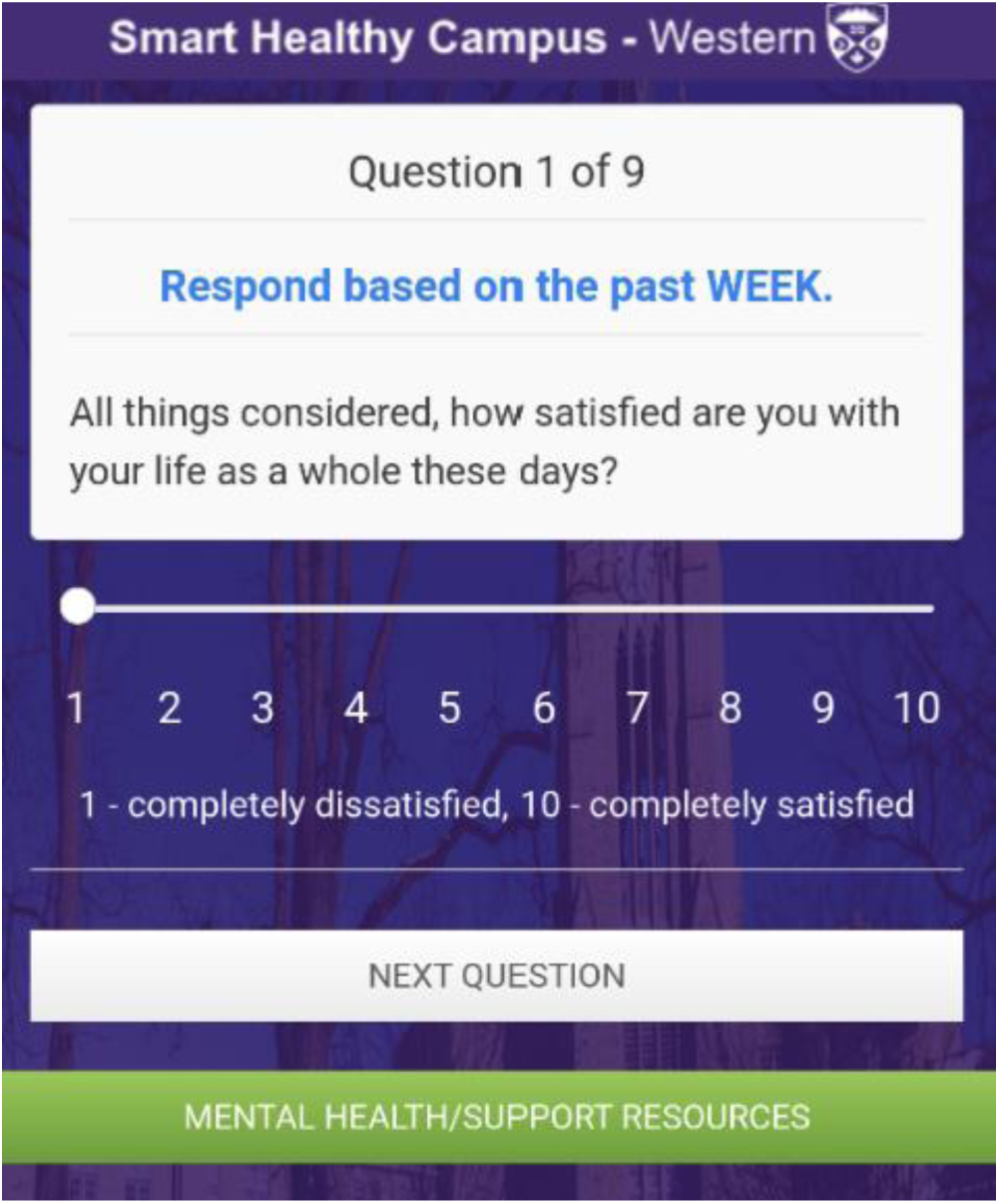
Image of the Smart Healthy Campus 2.0 app during a questionnaire.

## Methods

### Recruitment

Mass email recruitment was conducted. All Western University undergraduate students were invited over email to download the SHC 2.0 app in January 2021 and in March 2021 in order to participate in the study. The app was available on Google Play (Android) and the Apple App store. Sign-up information, including demographic information, was collected entirely in the app. To sign up for the SHC 2.0 app, participants had to view and accept the terms of the letter of information. At sign-up, they were also allowed to consent to providing their Twitter username (for tweet analysis) and to linking their SHC 2.0 app data with health records and psychological services records from campus health services. However, due to low consent rates, these sources of additional data were not used for SHC 2.0. 94 participants signed up for the app, and 86 participants both signed up and did at least one questionnaire in the app.

### Inclusion and Exclusion Criteria

The only inclusion criterion was that a participant was a full-time undergraduate student at Western University. Exclusion criteria were students enrolled in graduate or professional programs. Participants had to select their undergraduate program from a list in the SHC 2.0 app during the sign-up process.

### Study Design

SHC 2.0 was an Ecological Momentary Assessment-type app-based longitudinal cohort study with a repeated measures design. The SHC 2.0 app collected responses to a short questionnaire with a range of data from phone hardware potentially relevant to student mental health; responses and data were sent encrypted to our server when a participant submitted a questionnaire. The SHC 2.0 app also collected this same phone data in the background automatically on an hourly basis; participants could set this collection event to 1, 2, or 3 hours. This paper reports on the first 40 weeks (out of 52 weeks) of data collected for SHC 2.0; the majority of the study data was collected during this time. Participants could download and use the SHC 2.0 app and complete surveys as often or as little as they wanted, as there were no specific requirements on participation. As a result, compliance rates and missingness vary widely each day/week.

### Participant Compensation

The SHC 2.0 app included a points-based incentive system. Participants could accumulate points through app use and redeem 30000 points for a $5 Amazon gift card. Rewards were offered as the study progressed, and there were no limits on the total amount of gift cards redeemed. The first survey of the week, the weekly survey, would yield 3000 points. Daily surveys would provide 500 points, and background data collection events would provide 100. These point values were selected to prevent too much incentivization during a single week. Figure 2 shows an example of using the incentive system. It also allowed for the potential to add new rewards as the study progressed, although only the $5 Amazon gift cards were used during the study.

**Figure 2.**
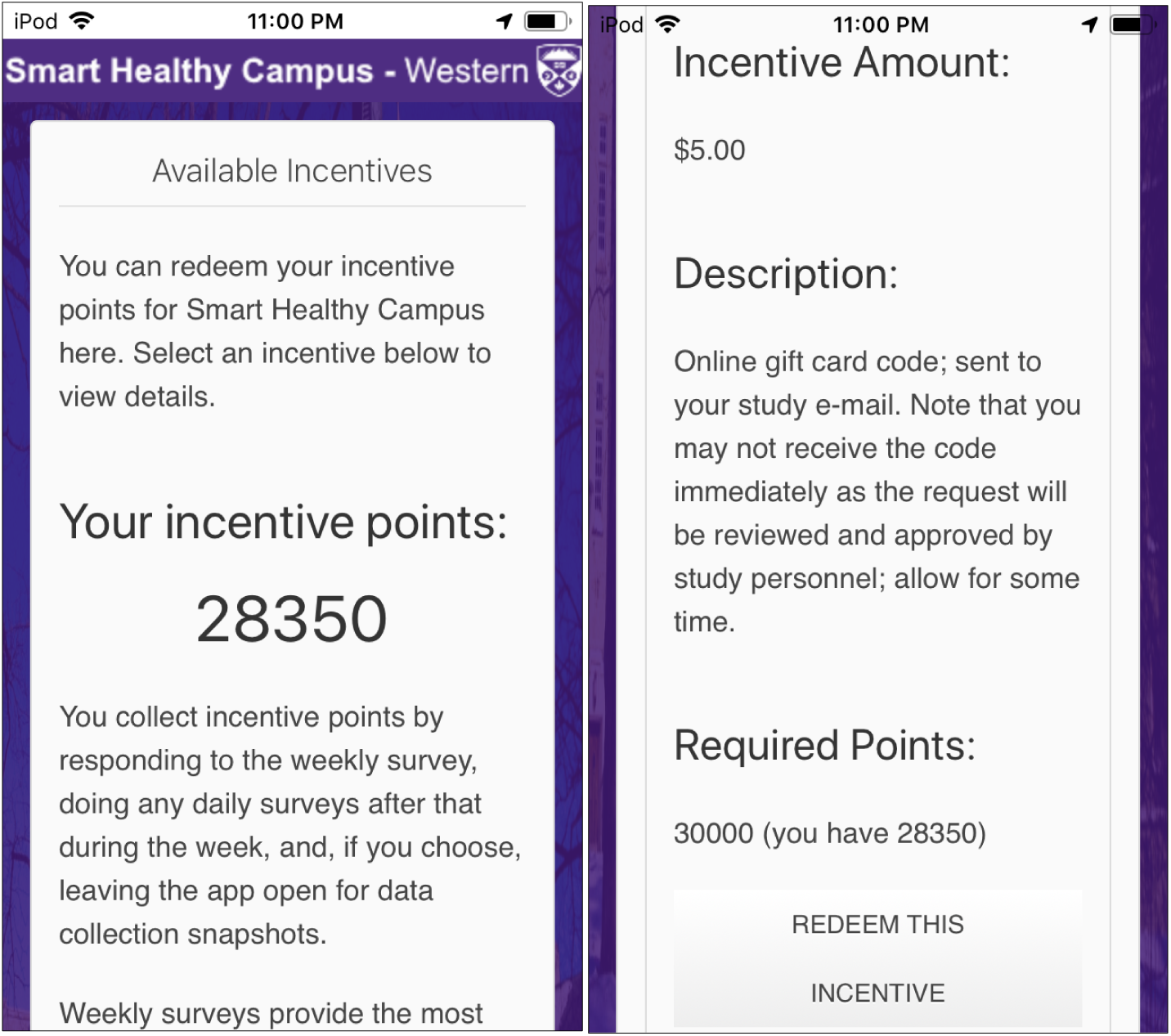
SHC 2.0 point-based incentive system screen (left image). The app has to be scrolled to see the entire screen (right image).

### Measures – Smart Healthy Campus 2.0 Questionnaire – Details

The SHC 2.0 app always makes a 9-item questionnaire available once per day. However, there are two variants to this questionnaire; the first is the weekly variant, which available during a new week starting Monday. The weekly variant was designed to capture the experience of the entire past week in case participants were infrequent responders. It would remain open as the first questionnaire to be completed for that week. As an example, if a participant waited until Friday or even Saturday or Sunday to provide a questionnaire response, the weekly variant would still be available then, since that would be the first questionnaire response of that week. If the weekly variant was completed, the daily variant was then available every day until the next Monday, including on Saturday and Sunday. These variants are very similar – only 2 of the 9 questions change and the text of the questions were edited to reflect either the daily or weekly focus. If a participant did not complete at least the weekly variant, then they did not provide any responses at all for that given week, although hourly background data may still have been collected by the SHC 2.0 app. Reminders to complete questionnaires were sent to participant devices via standard push notification manually by research personnel, usually at the beginning of the week, and occasionally throughout the week, but a set schedule for reminders was not followed. These notifications were generic reminders for all participants and did not change if a participant had completed a survey for that day.

Question topics from both variants cover a range of mental health-related measures relevant to students. For instance, two questions on resilience (weekly/daily) were included. Resilience is important for mental health, and is the ability to bounce back from, and adapt to difficult situations ^16^. One question on community connectedness was included (weekly/daily) and was of general interest to this study during typical in-person undergraduate studies, but the shift to online learning during the pandemic likely resulted in this having different but potentially interesting connotations.

### Measures – Smart Healthy Campus 2.0 Questionnaire, Weekly Variant

The weekly variant is almost the same as our pilot SHC 1.0 study questionnaire (which was also distributed weekly) and this was shown in the pilot SHC 1.0 study to significantly correlate with mental health domains based on validated full-length questionnaires ^13^. The first 7 questions are the same as the SHC 1.0 questionnaire. The last two new questions were considered for but disused in the original study; these were added here to increase data collection.

### Measures – Smart Healthy Campus 2.0 Questionnaire, Daily Variant

The daily variant was again based on the pilot SHC 1.0 questionnaire, and the content of the first 7 questions was the same, but they had more edits to clarify the questions were reflective of the day. Like the weekly variant, the last two new questions were considered for, but disused, in the initial SHC 1.0 pilot study and were again added here to increase data collection.

**Table 1.** Mental health domain coverage of the SHC 2.0 weekly and daily questionnaire variants. This table lists the domain of each of the 9 questions and its source. The two questionnaires are available in Multimedia Appendix 1.

| Weekly Variant<br>Mental Health<br>Domain | Source | Daily Variant Mental<br>Health Domain | Source |
| --- | --- | --- | --- |
| Life Satisfaction | 17 | Life satisfaction | 17 |
| Psychological well-being | 18 | Psychological well-being | 18 |
| Resilience | 19 | Resilience | 19 |
| Anxiety | 20 | Anxiety | 20 |
| Depression | 21 | Depression | 21 |
| Community<br>connectedness | 22 | Community<br>connectedness | 22 |
| Physical Activity | 23 | Anxiety | 21 |
| Anxiety | 21 | Psychological Distress | 24,25 |
| Resilience | 19 | Resilience | 19 |

### Digital Measures – Overview

From the SHC 2.0 app on the participant devices, 12 digital measures were selected for analysis in this paper (these data came almost exclusively from smartphones but tablets still were occasionally used). There were more than 12 measures collected via the SHC 2.0 app, but these 12 selected for analysis here had the greatest amount of data available across both Android and iOS platforms. A full list of digital measures that the SHC 2.0 app collected can be found in our paper on the EMAX platform which powered the SHC 2.0 app ^12^.

These digital measures were collected during two types of data collection “events”: Response and Background. Response events occurred whenever a participant responded to a survey. Background events occurred as a result of ongoing data collection, which happened on a 1, 2, or 3 hour basis (approximately). Participants could use a slider in the app to choose whether background data collection should occur approximately every 1, 2, or 3 hours. For both the Response and Background events, digital measure data were uploaded to our server. The Response event, unlike the Background event, also contained the response data to a weekly survey. Since Background events did not have this data, but it was needed for mixed model fits, an algorithm was run over all the Background event data to connect it to the most recent Response questionnaire result found before the Background event occurred.

### Digital Measures - Movement and Physical Activity

To determine associations between mental health questionnaire results and movement and physical activity, we examined 4 sources of data. First, we looked at total weekly step count. Step counts are commonly analyzed in mobile sensing studies ^26^. Second, we examined GPS locations visited. We define GPS locations visited as a sum of any significant changes in geographic location. Our SHC 2.0 app only detected GPS coordinates to two decimal places, so a “significant change” of at least 0.01 in the latitude/longitude of a coordinate is required to increase this sum. Third, we define floors walked as floors gone up + floors gone down over the past week. Fourth, a location “flag” was used (called “Campus Dist.” in the results section), and it could have three values: (1) on-campus, (2) in London, Canada and (3) outside of London, Canada.

### Digital Measures - Device usage

To determine associations between mental health questionnaire results and device usage, we examined 5 sources of data. First, we collected system uptime in milliseconds. We also collected system uptime (including deep sleep), in milliseconds (uptime + time the device was sleeping). Second, we obtained available RAM (available phone Random Access Memory) in bytes. Unlike CPU usage, available RAM (in bytes) is straightforward to collect across iOS/Android platforms. Third, we obtained a percentage of internal free space. Finally, we timed how long users had run the SHC 2.0 app on their device, in milliseconds.

### Digital Measures – Social/life activity indicators

Contacts and calendars are data sources that may be indicative of social or life activity and they were straightforward to access across platforms. The sum of calendar events were collected. These events may include items like reminders for physical activity, events, and appointments. A sum of all contact counts at each sample was also obtained. Additionally, a basic algorithm to check for family-related names (or containing family-related names) in contacts was run to collect a sum of these.

### Digital Measures - Additional item: Number of apps installed

One additional data source was included in this analysis and considered a digital measure. This was a percentage of popular installed apps from a known list of 40. However, we did not split these by category due to the small list of 40.

### Statistical Analysis

The focus of the statistical analysis in this paper is on the relationship between phone digital measures and the SHC 2.0 questionnaire variants. The total score of the SHC 2.0 questionnaire variants are examined (some items were recoded to make them suitable for a total sum), along with scores from individual questions, which are representative of various mental health domains ^13^ (see Methods). SHC 2.0 required participants to answer all questions since the questionnaires were short. R 4.1.1 “Kick Things” and RStudio were used for all statistics. All mixed linear models shown were constructed using the lmerTest library ^27^ which provides p-values for effects using the Satterthwaite degrees of freedom method ^27^. sjPlot was used for coefficient graphs and ggplot2 was used for line graphs. This approach to mixed linear models with EMA data and the same R packages were previously used by Huckins et al. ^28^ and Mack et al. ^29^ in their study on student mental health and the impact of COVID-19 at Dartmouth College.

With regards to building the mixed linear models, z-score standardization was used on digital measure items with R’s scale(…) function as values could vary considerably. For each mental health questionnaire (DV), a first model was fit with all 12 previously discussed digital measure items as fixed-effects. The participants were the random-effect. Then, all the digital measure items that lmerTest did not report significance for were removed, and a second, final model was fit with only the significant digital measure items (backward elimination). The reason for fitting and only using a second model with backward elimination was that removing those insignificant terms sometimes greatly increased the amount of complete case digital measure samples available for use in the lmerTest models. It also removed digital measure items with weaker (although in the first model, initially significant) p-values.

#### Data Inclusion

Highly correlated mixed model parameters can make each other non-significant even if they are predictive. There were many significant correlations between the parameters used for these models, however, the majority were very weak or weak based on Spearman coefficients (0.0-0.20). Some coefficients did enter moderate territory in the range of 0.4 to 0.6. Additionally, there was a strong correlation between system uptime, and system uptime with deep sleep, although we argue this was acceptable as those likely better highlighted differences between device use and potential idle or sleep time for participants; additionally, more than one model resulted in significance for only one of these parameters, while others produced significance for both, suggesting the correlation was not important. We argue these correlations in our 12 selected digital measure items, most of them being very weak or weak, were not detrimental to the mixed model fits as the 12 selected items were the most available across participants and platforms (iOS, Android), and thus best representative of lifestyle and behaviors during this time.

Since backward elimination was applied to remove insignificant digital measure items from final models, sample sizes and the associated number of complete case digital measure sample observations used for the model fits may vary, so they are shown with the associated model in the results section.

## Results

### Overview

Our main results consist of 17 mixed linear models. Note that the mixed model “Measures” (fixed-effects) such as “RAM” or “GPS Locations Visited” are described in the “Digital Measures” sections under Methods. There are two rounds of fits for the 17 mixed models. The second, revised models are based on backward elimination of the first in order to produce only the significant fixed-effects (device digital measures) that might predict scores for SHC 2.0 weekly/daily questionnaires and also for individual questions. This backward elimination process was used to increase the complete case count - samples that contained data for all 12 digital measures selected. Individual questions are examined here in addition to the total questionnaire score, as on their own, they were designed to capture various domains of mental health (e.g. depression, anxiety) ^13^ (discussed in Methods). The goal of the mixed linear model analysis is that by knowing the measurable behaviors/lifestyle associated with positive mental health, it should become easier to design traditional or computer-related interventions to direct participants to those associated behaviors/lifestyle. Another goal is that these models might be integrated into future apps that directly provide interventions or predictions, that might be triggered by these models or similar ones.

First, the demographics of the 94 participants in SHC 2.0 are presented. Then, we present plots of selected mean scores from our SHC 2.0 weekly questionnaire with key pandemic event markers. Lastly, the results of the 17 mixed linear model fits are shown between the 12 selected phone digital measures and A) the SHC 2.0 daily questionnaire total score and individual questions, and B) the SHC 2.0 weekly questionnaire total score and individual questions. The results of the daily questionnaire model fits are presented to start as more data was collected daily.

### Demographics

Table 2 below presents the demographic information of the 94 participants in SHC 2.0. Participants primarily identified as male and living in off-campus housing.

**Table 2.**
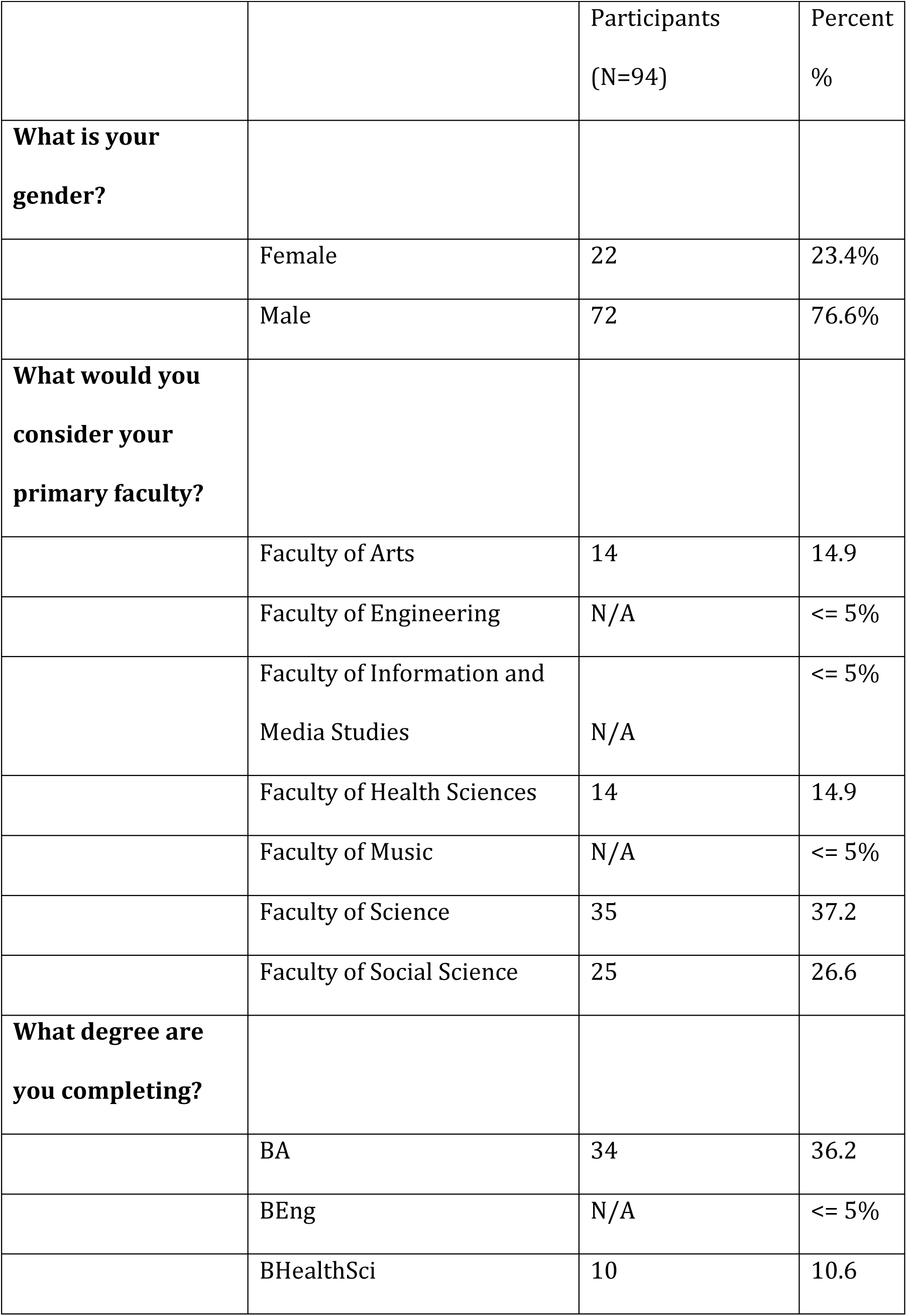
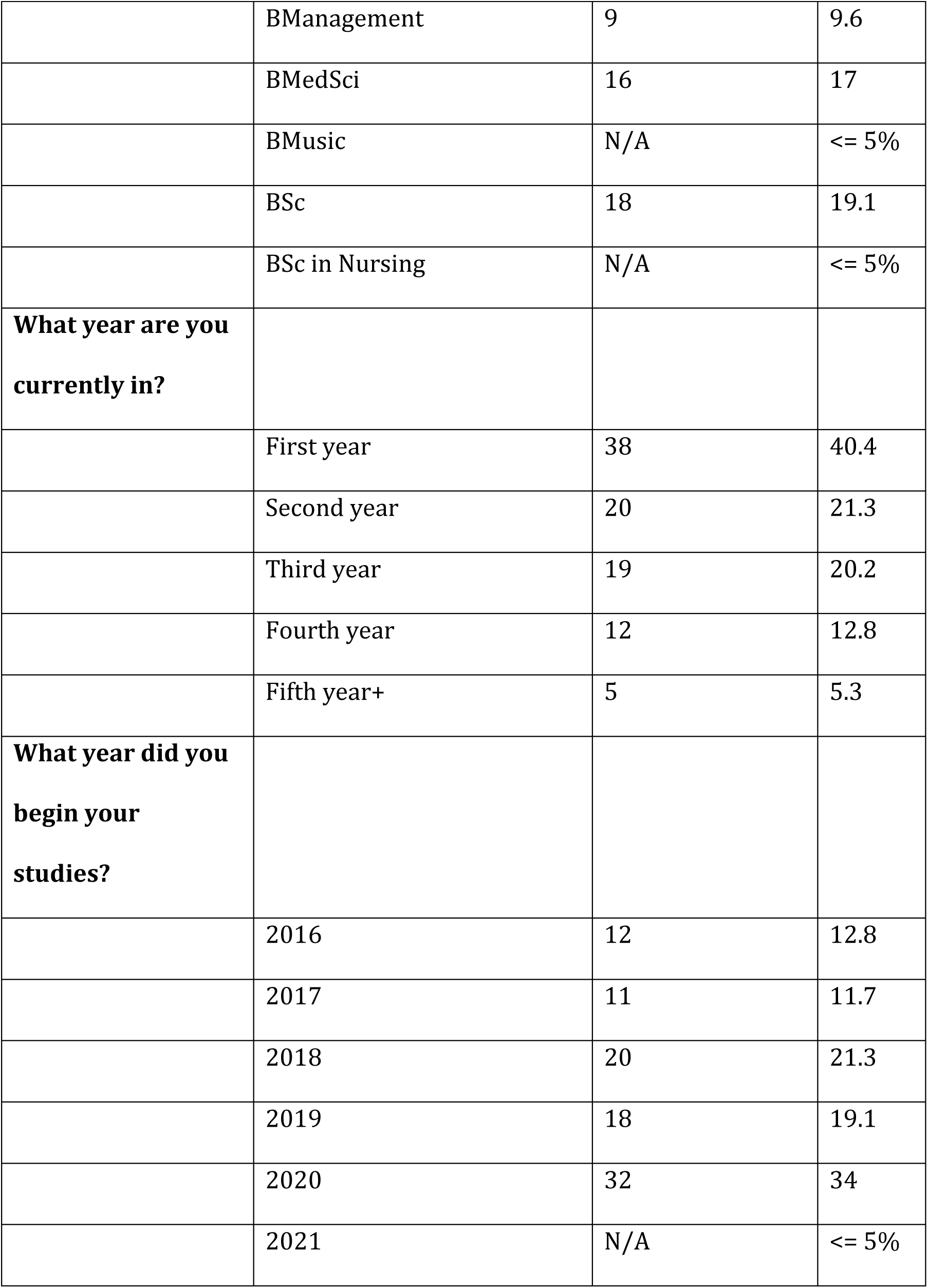
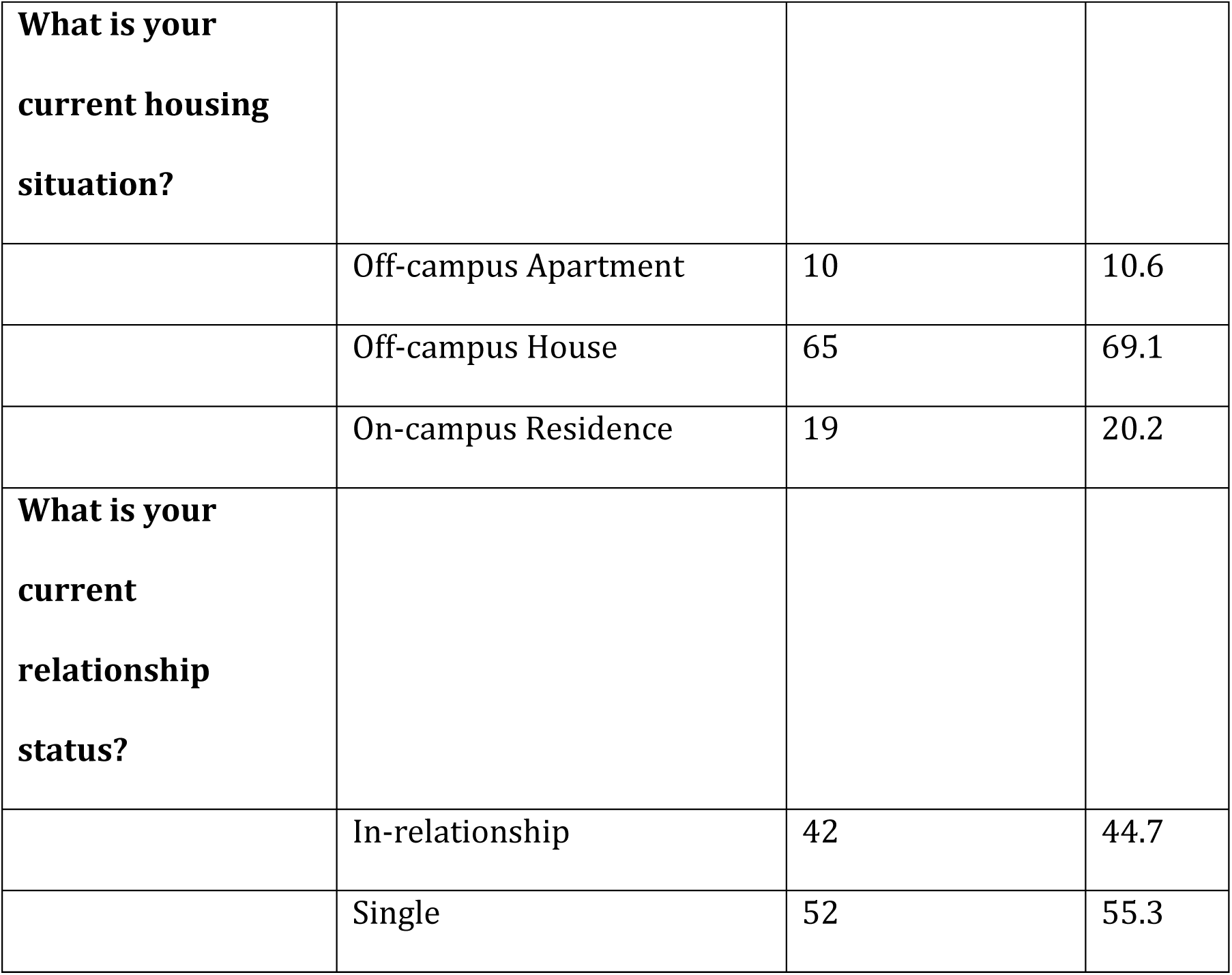
Participant demographics at baseline. As questions can be skipped, sometimes the cells for each item may not necessarily add to 100%.

|  |  | Participants<br>(N=94) | Percent<br>% |
| --- | --- | --- | --- |
| **What is your gender?** |  |  |  |
|  | Female | 22 | 23.4% |
|  | Male | 72 | 76.6% |
| **What would you consider your primary faculty?** |  |  |  |
|  | Faculty of Arts | 14 | 14.9 |
|  | Faculty of Engineering | N/A | <= 5% |
|  | Faculty of Information and Media Studies | N/A | <= 5% |
|  | Faculty of Health Sciences | 14 | 14.9 |
|  | Faculty of Music | N/A | <= 5% |
|  | Faculty of Science | 35 | 37.2 |
|  | Faculty of Social Science | 25 | 26.6 |
| **What degree are you completing?** |  |  |  |
|  | BA | 34 | 36.2 |
|  | BEng | N/A | <= 5% |
|  | BHealthSci | 10 | 10.6 |
|  | BManagement | 9 | 9.6 |
|  | BMedSci | 16 | 17 |
|  | BMusic | N/A | <= 5% |
|  | BSc | 18 | 19.1 |
|  | BSc in Nursing | N/A | <= 5% |
| **What year are you currently in?** |  |  |  |
|  | First year | 38 | 40.4 |
|  | Second year | 20 | 21.3 |
|  | Third year | 19 | 20.2 |
|  | Fourth year | 12 | 12.8 |
|  | Fifth year+ | 5 | 5.3 |
| **What year did you begin your studies?** |  |  |  |
|  | 2016 | 12 | 12.8 |
|  | 2017 | 11 | 11.7 |
|  | 2018 | 20 | 21.3 |
|  | 2019 | 18 | 19.1 |
|  | 2020 | 32 | 34 |
|  | 2021 | N/A | <= 5% |
| **What is your current housing situation?** |  |  |  |
|  | Off-campus Apartment | 10 | 10.6 |
|  | Off-campus House | 65 | 69.1 |
|  | On-campus Residence | 19 | 20.2 |
| **What is your current relationship status?** |  |  |  |
|  | In-relationship | 42 | 44.7 |
|  | Single | 52 | 55.3 |

### Pandemic events and selected mean scores of SHC 2.0 weekly questionnaire

Although SHC 2.0 was not specifically designed to run during the pandemic, it ended up being launched during COVID-19 and ran for a significant amount of time during the pandemic, as the app’s data collection and questionnaire remained relevant. We provide visualizations of select mental health measures overlayed by key provincial lockdown measures in Ontario below. The mean scores of the sum of the SHC 2.0 weekly questionnaire, along with means of selected individual questions from the weekly variant, are shown in Figures 3 and 4. Important dates related to the COVID-19 pandemic are shown in Table 3.

**Figure 3.**
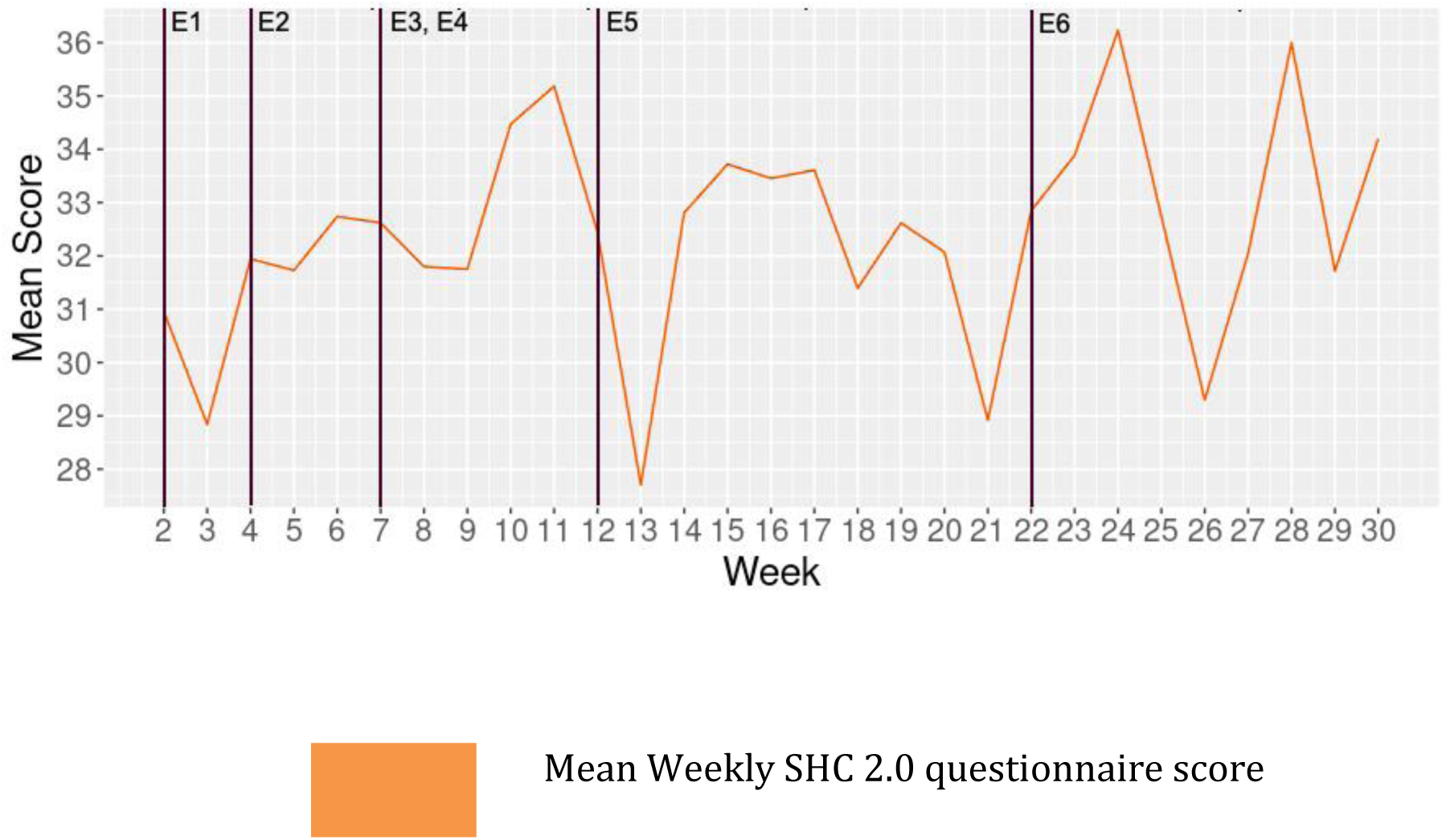
SHC 2.0 mean weekly questionnaire score over study weeks during COVID-19. The weekly score is representative of a broad overview of participant mental health (Life satisfaction, psychological well-being, resilience, anxiety, depression, and community connectedness). Higher scores are representative of a more positive mean overview (ie. better).

**Figure 4.**
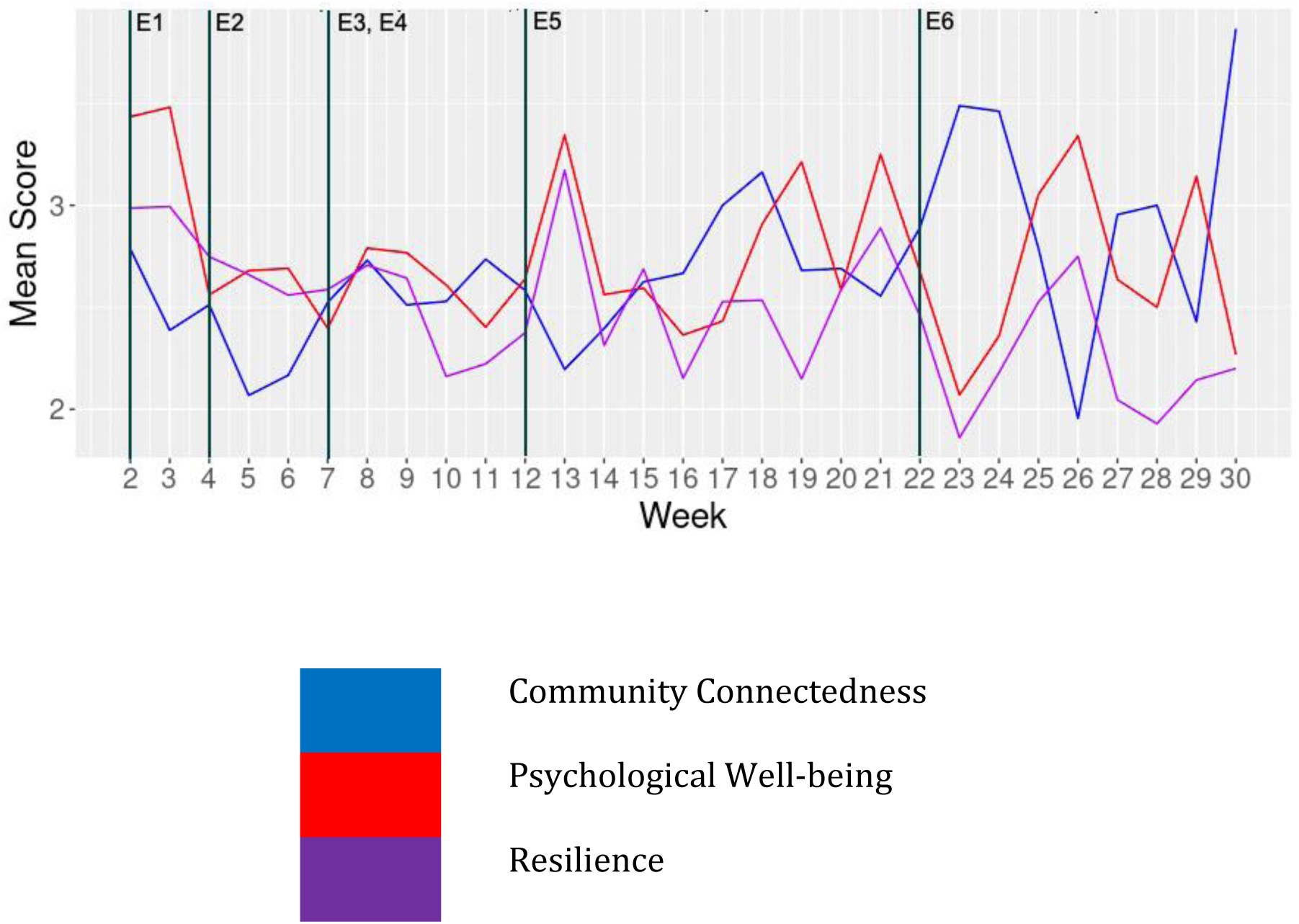
Selected measures from SHC 2.0 over study weeks during the COVID-19 pandemic: community connectedness, psychological well-being, and resilience. Higher scores can be considered positive (ie. better).

**Table 3.**
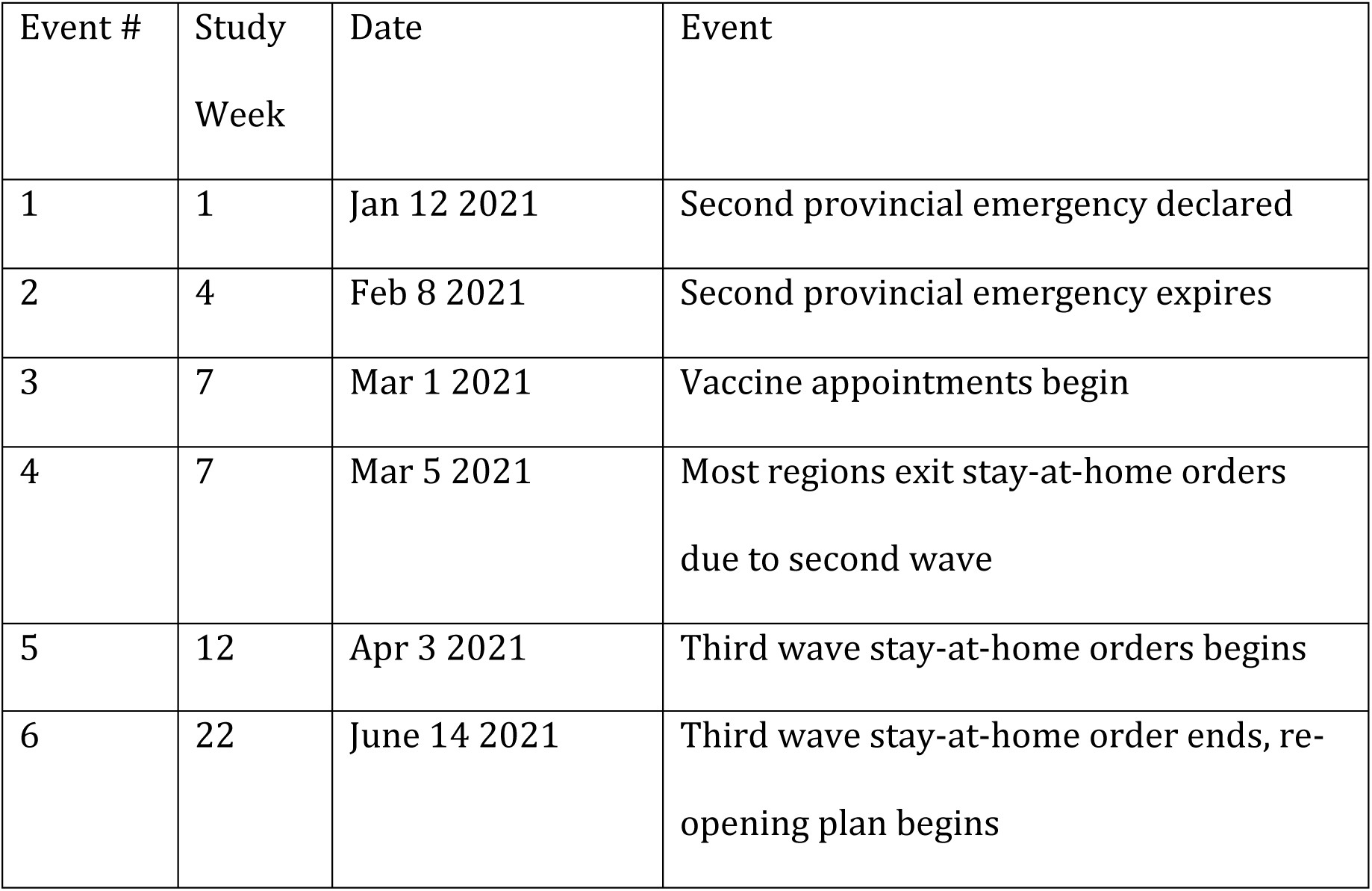
Key events related to the SHC 2.0 study and COVID-19 during January 2021-June 2021. These correspond to the event lines (E1-E6) in Figure 3 and 4.

| Event # | Study Week | Date | Event |
| --- | --- | --- | --- |
| 1 | 1 | Jan 12 2021 | Second provincial emergency declared |
| 2 | 4 | Feb 8 2021 | Second provincial emergency expires |
| 3 | 7 | Mar 1 2021 | Vaccine appointments begin |
| 4 | 7 | Mar 5 2021 | Most regions exit stay-at-home orders due to second wave |
| 5 | 12 | Apr 3 2021 | Third wave stay-at-home orders begins |
| 6 | 22 | June 14 2021 | Third wave stay-at-home order ends, re-opening plan begins |

#### Interpretation of coefficient plots

This section explains how to interpret the coefficient plots of the mixed models below. Every plot shows the amount of change associated with the dependent variable (questionnaire measure) as there is a +1 unit change in the digital measure item (independent variable, y-axis). Note that digital measure items were z-score standardized due to different very scales, so a +1 unit change is a +1 standard deviation from the mean. Dependent variables were not standardized for future prediction purposes. Essentially, an association can be positive or negative, and the value on the x-axis “Estimates” for any predictor on the y-axis would be added to the dependent variable as there is a +1 unit change for that y-axis predictor.

### Mixed linear models of SHC 2.0 daily questionnaire variant

In this section, the mixed linear model fits for the measures of the SHC 2.0 daily questionnaire are presented. In the second round of model fits, significant fixed effects were found for all the daily variant’s individual questions and for the total questionnaire sum.

#### Regressing daily questions 1-3 on device digital measures

**Table 4.**
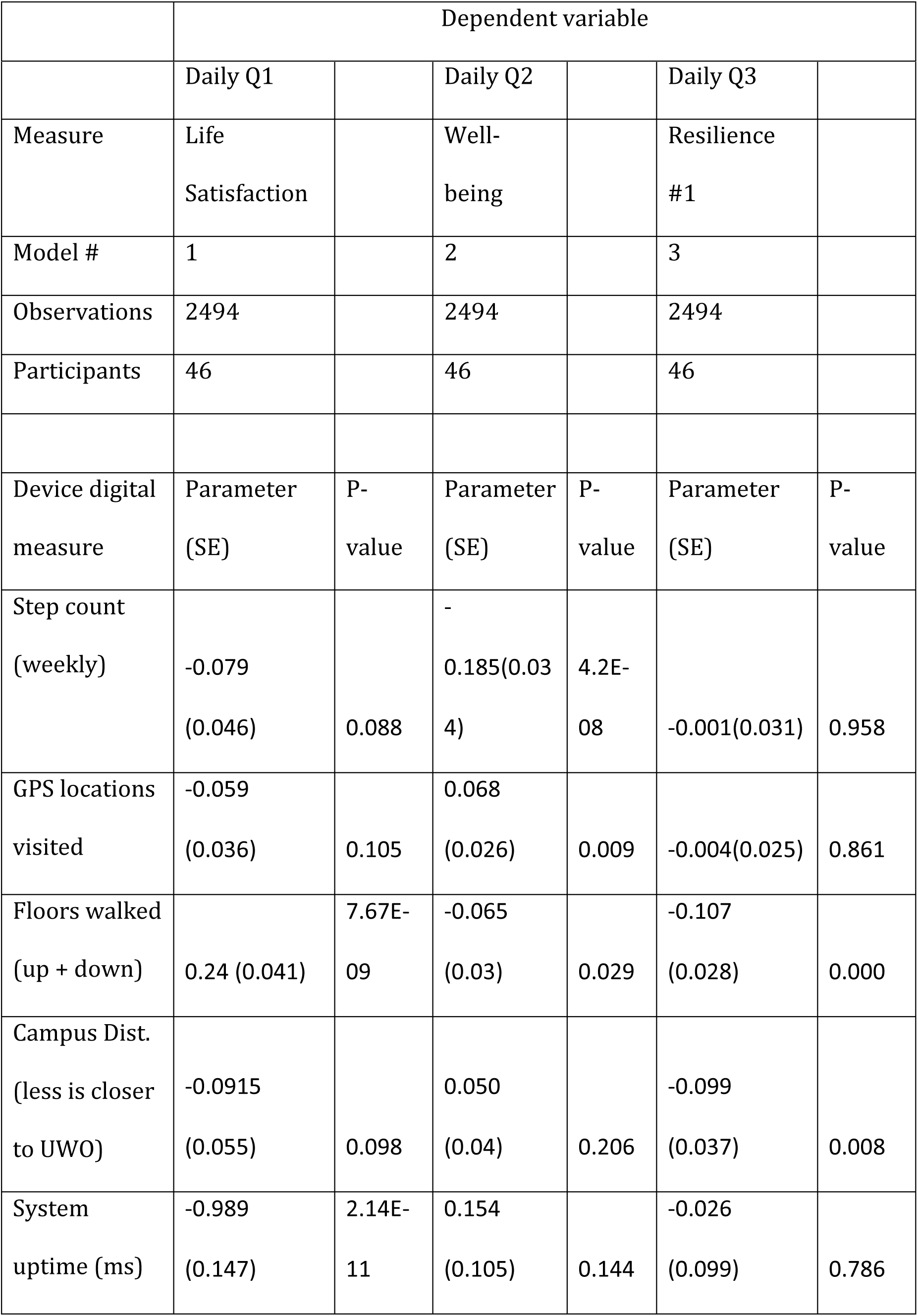
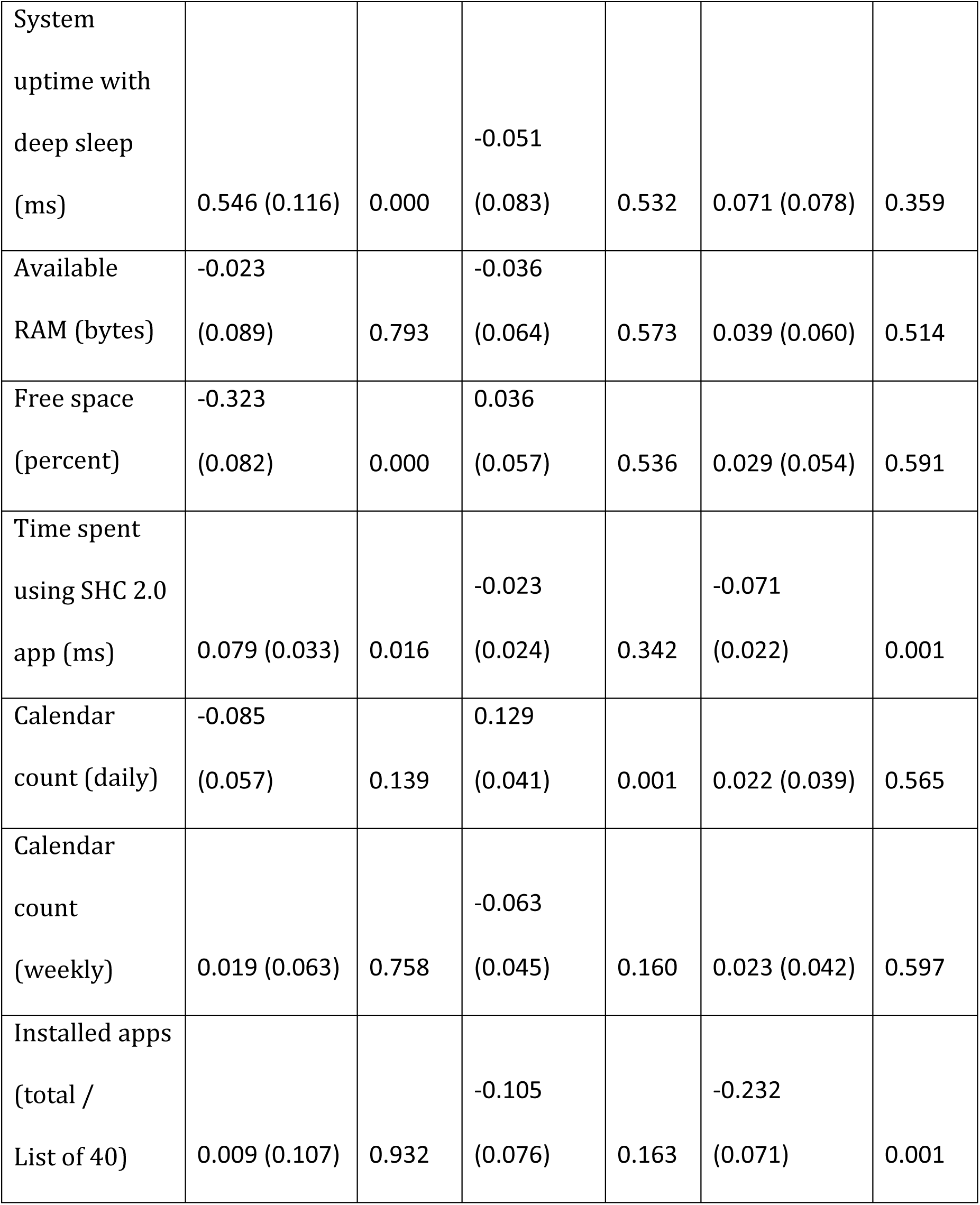
First round of mixed linear models for daily life satisfaction, daily psychological well-being, and daily resilience.

|  |  |  |  |  |  |  |
| --- | --- | --- | --- | --- | --- | --- |
|  | Daily Q1 |  | Daily Q2 |  | Daily Q3 |  |
| Measure | Life Satisfaction |  | Well-being |  | Resilience #1 |  |
| Model # | 1 |  | 2 |  | 3 |  |
| Observations | 2494 |  | 2494 |  | 2494 |  |
| Participants | 46 |  | 46 |  | 46 |  |
| Device digital measure | Parameter (SE) | P-value | Parameter (SE) | P-value | Parameter (SE) | P-value |
| Step count (weekly) | -0.079<br>(0.046) | 0.088 | -<br>0.185(0.034) | 4.2E-08 | -0.001(0.031) | 0.958 |
| GPS locations visited | -0.059<br>(0.036) | 0.105 | 0.068<br>(0.026) | 0.009 | -0.004(0.025) | 0.861 |
| Floors walked (up + down) | 0.24 (0.041) | 7.67E-09 | -0.065<br>(0.03) | 0.029 | -0.107<br>(0.028) | 0.000 |
| Campus Dist. (less is closer to UWO) | -0.0915<br>(0.055) | 0.098 | 0.050<br>(0.04) | 0.206 | -0.099<br>(0.037) | 0.008 |
| System uptime (ms) | -0.989<br>(0.147) | 2.14E-11 | 0.154<br>(0.105) | 0.144 | -0.026<br>(0.099) | 0.786 |
| System<br>uptime with<br>deep sleep<br>(ms) | 0.546 (0.116) | 0.000 | -0.051<br>(0.083) | 0.532 | 0.071 (0.078) | 0.359 |
| Available<br>RAM (bytes) | -0.023<br>(0.089) | 0.793 | -0.036<br>(0.064) | 0.573 | 0.039 (0.060) | 0.514 |
| Free space<br>(percent) | -0.323<br>(0.082) | 0.000 | 0.036<br>(0.057) | 0.536 | 0.029 (0.054) | 0.591 |
| Time spent<br>using SHC 2.0<br>app (ms) | 0.079 (0.033) | 0.016 | -0.023<br>(0.024) | 0.342 | -0.071<br>(0.022) | 0.001 |
| Calendar<br>count (daily) | -0.085<br>(0.057) | 0.139 | 0.129<br>(0.041) | 0.001 | 0.022 (0.039) | 0.565 |
| Calendar<br>count<br>(weekly) | 0.019 (0.063) | 0.758 | -0.063<br>(0.045) | 0.160 | 0.023 (0.042) | 0.597 |
| Installed apps<br>(total /<br>List of 40) | 0.009 (0.107) | 0.932 | -0.105<br>(0.076) | 0.163 | -0.232<br>(0.071) | 0.001 |

#### Revised Models #1-3 using significant parameters only, with coefficient plots

**Table 5.** Revised second round (using only significant digital measure items) of mixed linear models for daily life satisfaction, daily psychological well-being, and daily resilience.

|  | Dependent variable |  |  |
| --- | --- | --- | --- |
|  | Life Satisfaction | Well-being | Resilience #1 |
| Revised Model # | 1 | 2 | 3 |
| Observations | 3356 | 3376 | 2514 |
| Participants | 58 | 58 | 46 |
| Device digital measure | Parameter (SE) | Parameter (SE) | Parameter (SE) |
| Step count<br>(weekly) |  | -0.189 (0.025) |  |
| GPS locations visited |  | 0.046 (0.022) |  |
| Floors walked (up +<br>down) | 0.115 (0.031) |  | -0.105 (0.022) |
| Campus Dist. (less is<br>closer to UWO) |  |  | -0.108 (0.034) |
| System uptime (ms) | -0.33 (0.058) |  |  |
| Free space<br>(percent) | -0.212 (0.079) |  |  |
| Time spent using SHC<br>2.0 app (ms) |  |  | -0.071 (0.020) |
| Calendar count (daily) |  | 0.105 (0.034) |  |
| Installed apps (total /<br>List of 40) |  |  | -0.226 (0.070) |

**Figure 5.**
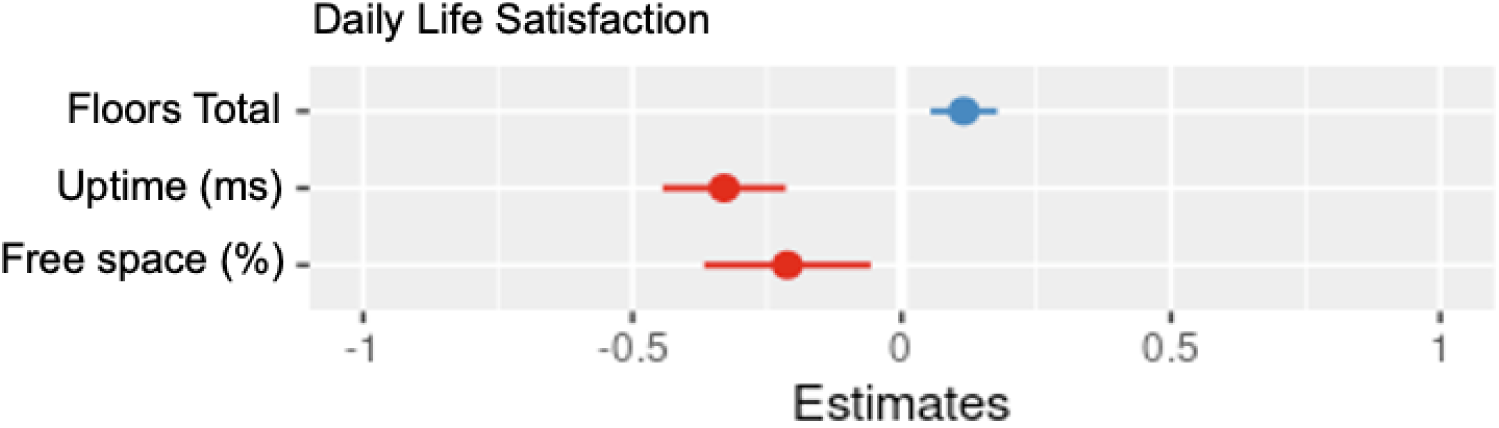
Coefficient plot for parameters of Model #1 – Daily Life Satisfaction.

System uptime had a negative association with life satisfaction. There was also a negative association with free space. This suggests less device usage may be associated with increased life satisfaction. There also appeared to be a positive association with floors walked. Based on this model, physical activity did not appear to be associated with increased life satisfaction.

**Figure 6.**
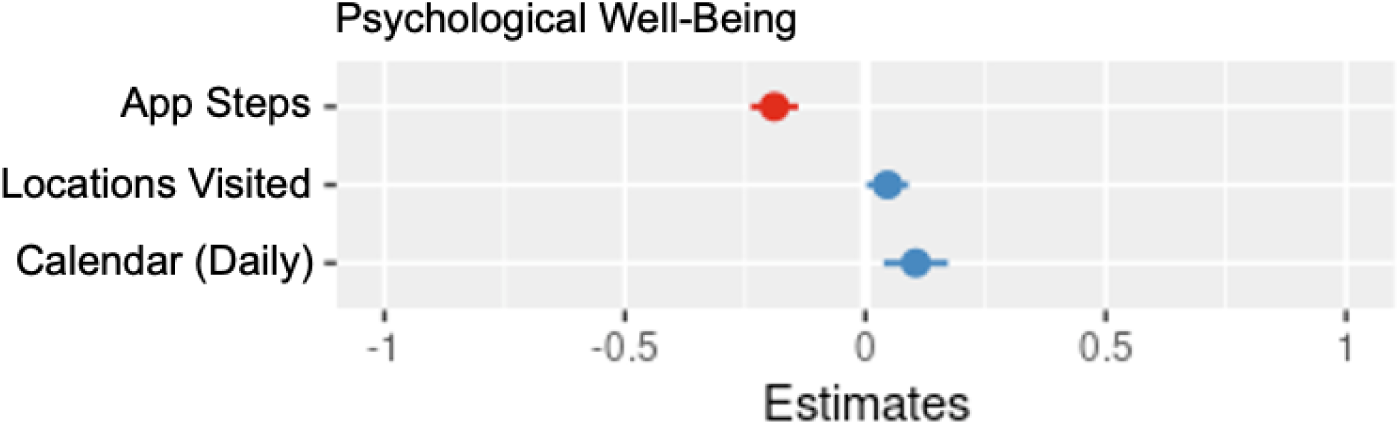
Coefficient plot for parameters of Model #2 – Daily Psychological Well-being.

Surprisingly, app steps had a negative association with daily psychological well-being. There were positive associations with locations visited (to an extent) and daily calendar counts, suggesting more life activity was related to psychological well-being.

**Figure 7.**
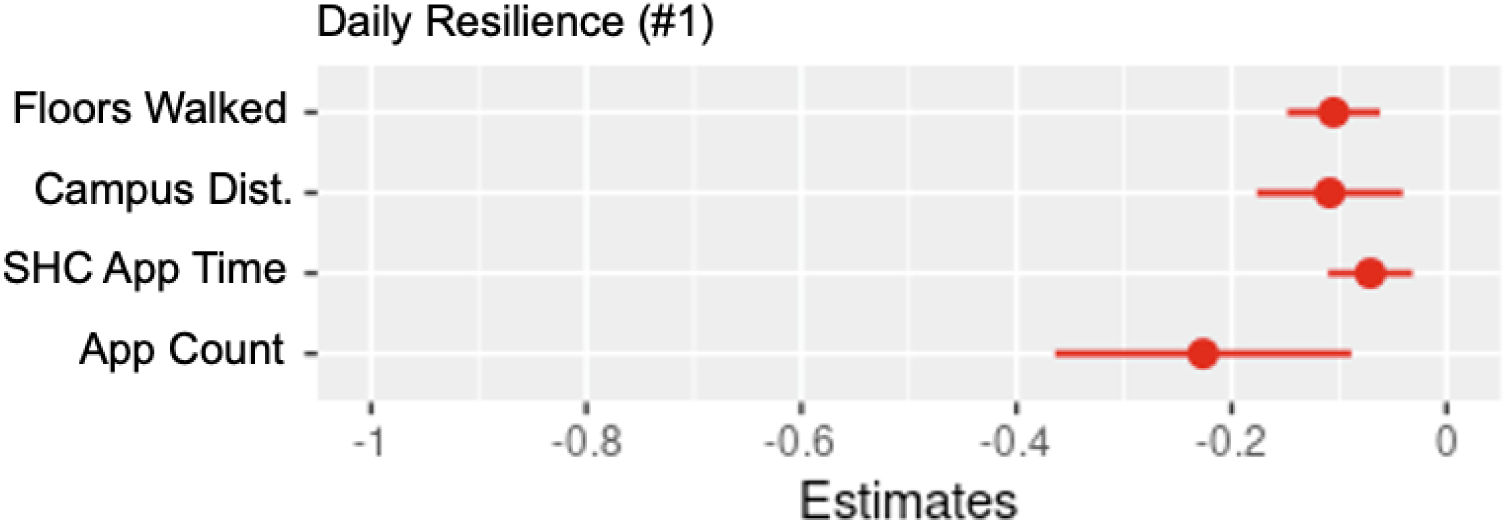
Coefficient plot for parameters of Model #3 – Daily Resilience Measure #1.

Floors walked, device usage via SHC 2.0 total app use time, and installed apps all had negative associations with the first daily measure of resilience. There also was a positive association with proximity to campus.

#### Regressing daily questions 4-6 on device digital measures

Table 6 shows the results of three mixed linear models relating device digital measures to a first daily measure of anxiety, a daily measure of depression, and a daily measure of community connectedness.

**Table 6.** First round of mixed linear models for daily anxiety, daily depression, and daily community connectedness.

|  | Dependent variable |  |  |  |  |  |
| --- | --- | --- | --- | --- | --- | --- |
|  | Daily Q4 |  | Daily Q5 |  | Daily Q6 |  |
| Measure | Anxiety #1 |  | Depression |  | Comm.<br>Connected. |  |
| Model # | 4 |  | 5 |  | 6 |  |
| Observations | 2494 |  | 2494 |  | 2494 |  |
| Participants | 46 |  | 46 |  | 46 |  |
| Device digital<br>measure | Parameter<br>(SE) | P-value | Parameter<br>(SE) | P-value | Parameter<br>(SE) | P-value |
| Step count<br>(weekly) | -0.04(0.021) | 0.069 | 0.061<br>(0.016) | 0.000 | 0.054 (0.028) | 0.057 |
| GPS locations<br>visited | 0.039 (0.017) | 0.022 | 0.026<br>(0.013) | 0.049 | 0.097 (0.022) | 0.000 |
| Floors walked<br>(up + down) | -0.051<br>(0.019) | 0.007 | -0.047<br>(0.014) | 0.001 | 0.0041<br>(0.025) | 0.871 |
| Campus Dist.<br>(less is closer<br>to UWO) | -0.022<br>(0.025) | 0.382 | 0.050<br>(0.019) | 0.011 | -0.127<br>(0.034) | 0.000 |
| System<br>uptime (ms) | 0.237 (0.068) | 0.000 | 0.254<br>(0.052) | 0.000 | 0.218 (0.090) | 0.017 |
| System<br>uptime with<br>deep sleep<br>(ms) | -0.067<br>(0.053) | 0.214 | -0.193<br>(0.042) | 0.000 | -0.208<br>(0.072) | 0.004 |
| Available<br>RAM (bytes) | 0.023 (0.041) | 0.564 | -0.008<br>(0.031) | 0.789 | -0.0545<br>(0.054) | 0.321 |
| Free space<br>(percent) | -0.062<br>(0.038) | 0.105 | 0.021<br>(0.031) | 0.498 | 0.072 (0.051) | 0.157 |
| Time spent<br>using SHC 2.0<br>app (ms) | 0.015 (0.015) | 0.328 | -0.013<br>(0.011) | 0.276 | -0.035<br>(0.020) | 0.085 |
| Calendar<br>count (daily) | 0.011 (0.026) | 0.686 | 0.006<br>(0.021) | 0.771 | -0.061<br>(0.036) | 0.084 |
| Calendar<br>count<br>(weekly) | -0.015<br>(0.029) | 0.622 | -0.016<br>(0.022) | 0.483 | 0.104 (0.039) | 0.008 |
| Installed apps<br>(total /<br>List of 40) | -0.148<br>(0.049) | 0.003 | -0.045<br>(0.039) | 0.255 | -0.225<br>(0.066) | 0.000 |

#### Revised Models #4-6 using significant parameters only, with coefficient plots

**Table 7.**
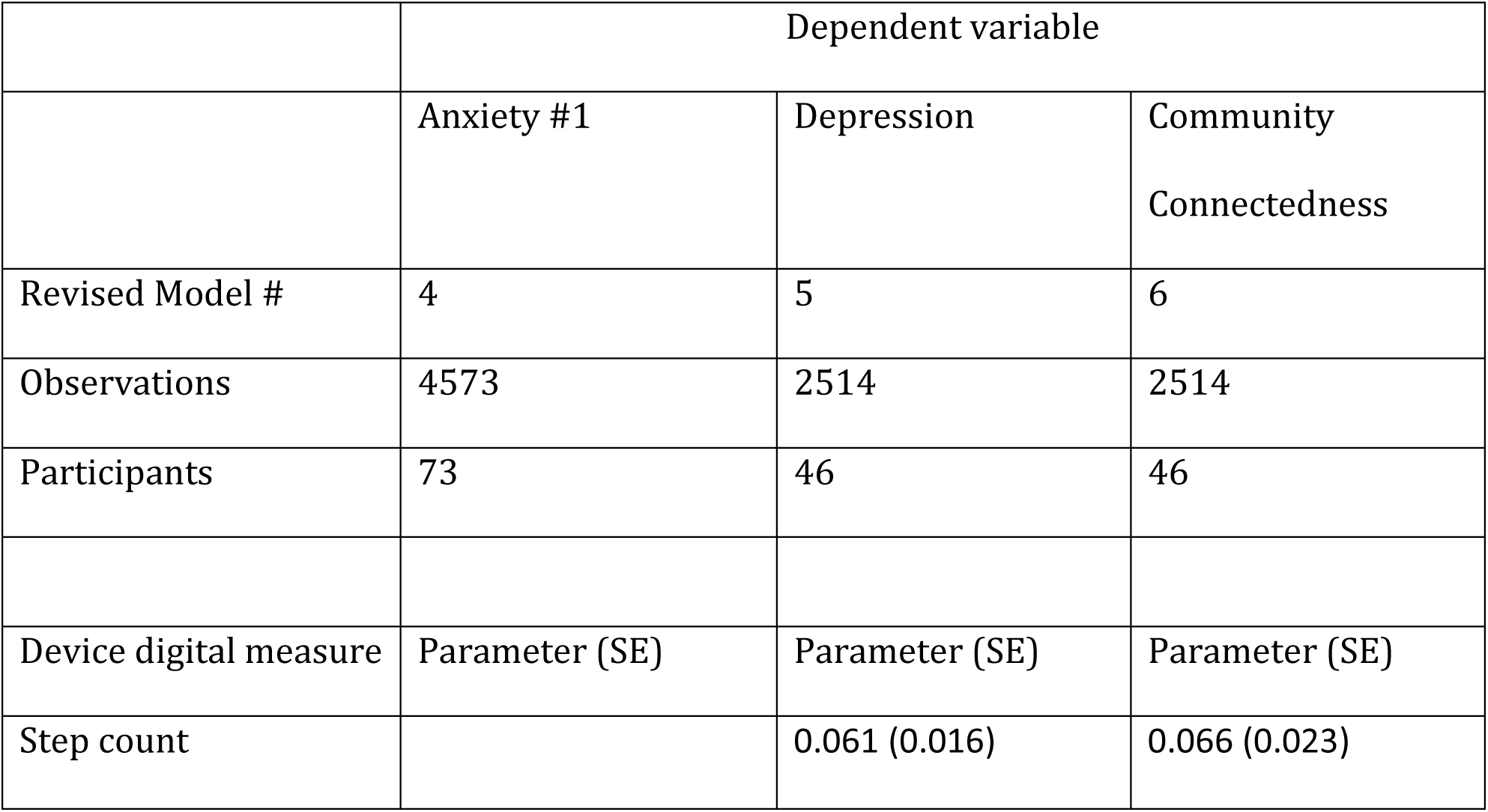
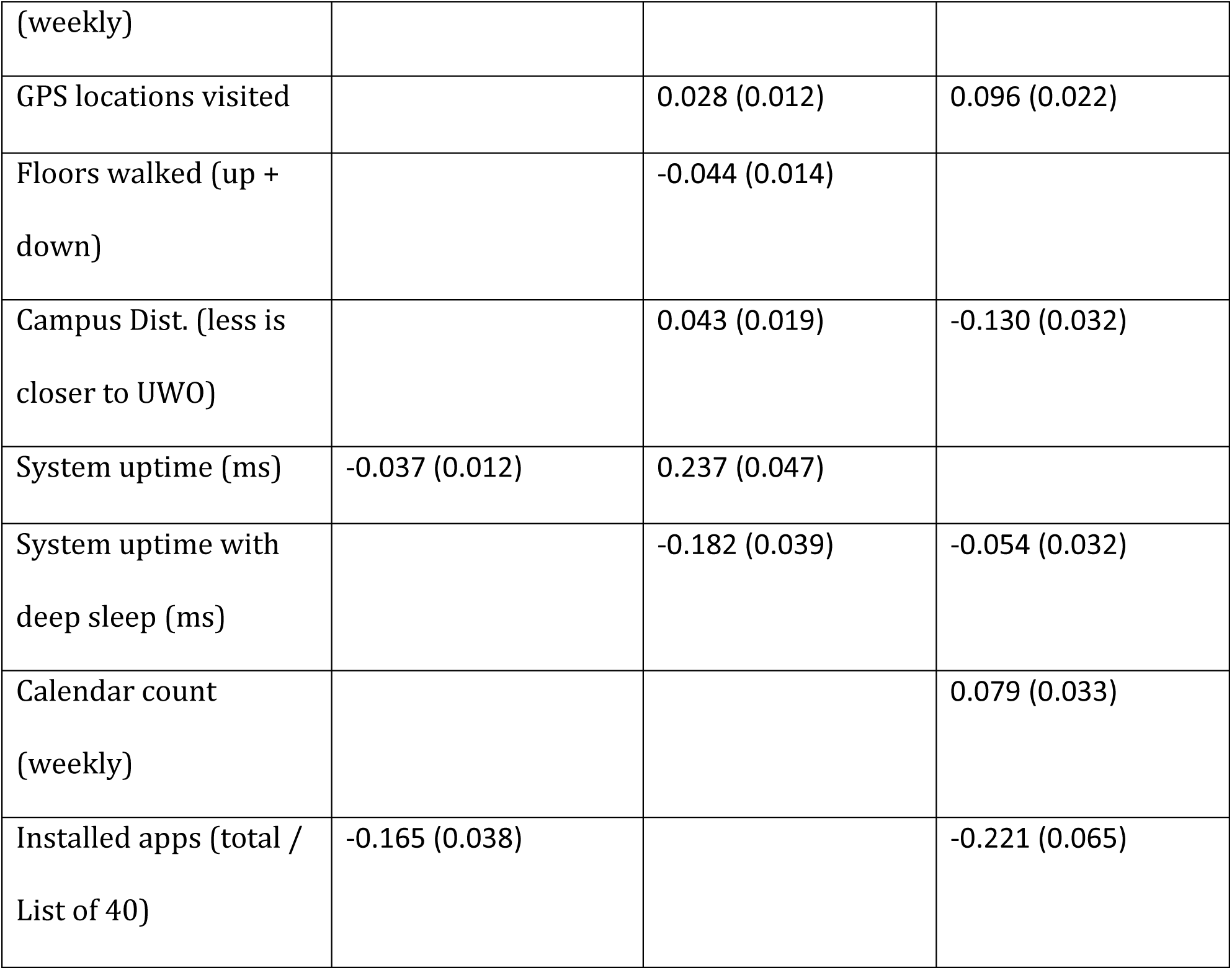
Revised second round (using only significant digital measure items) of mixed linear models for daily anxiety, daily depression, and daily community connectedness.

**Figure 8.**
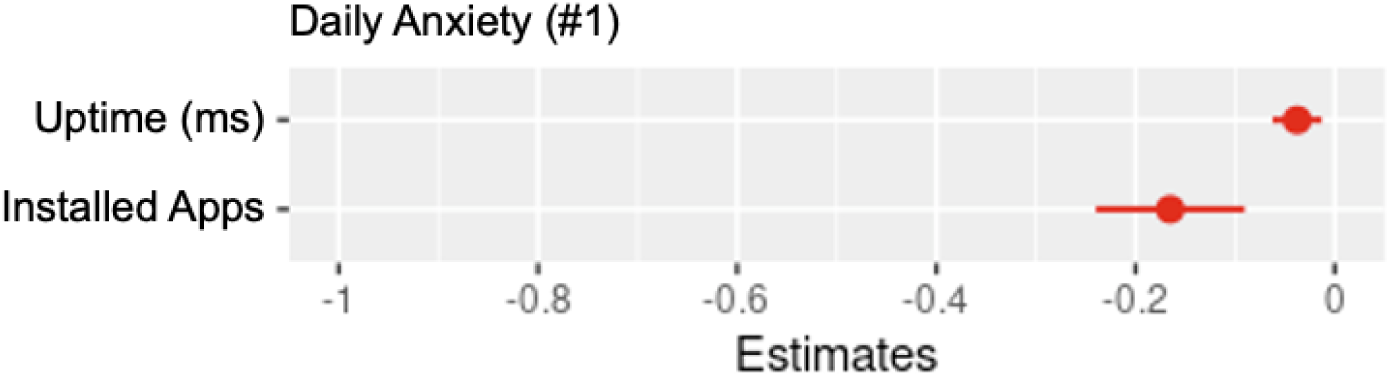
Coefficient plot for parameters of Model #4 – daily anxiety measure #1.

Uptime had a small negative association with the first measure of daily anxiety. There was a more significant negative association with the number of installed apps.

**Figure 9.**
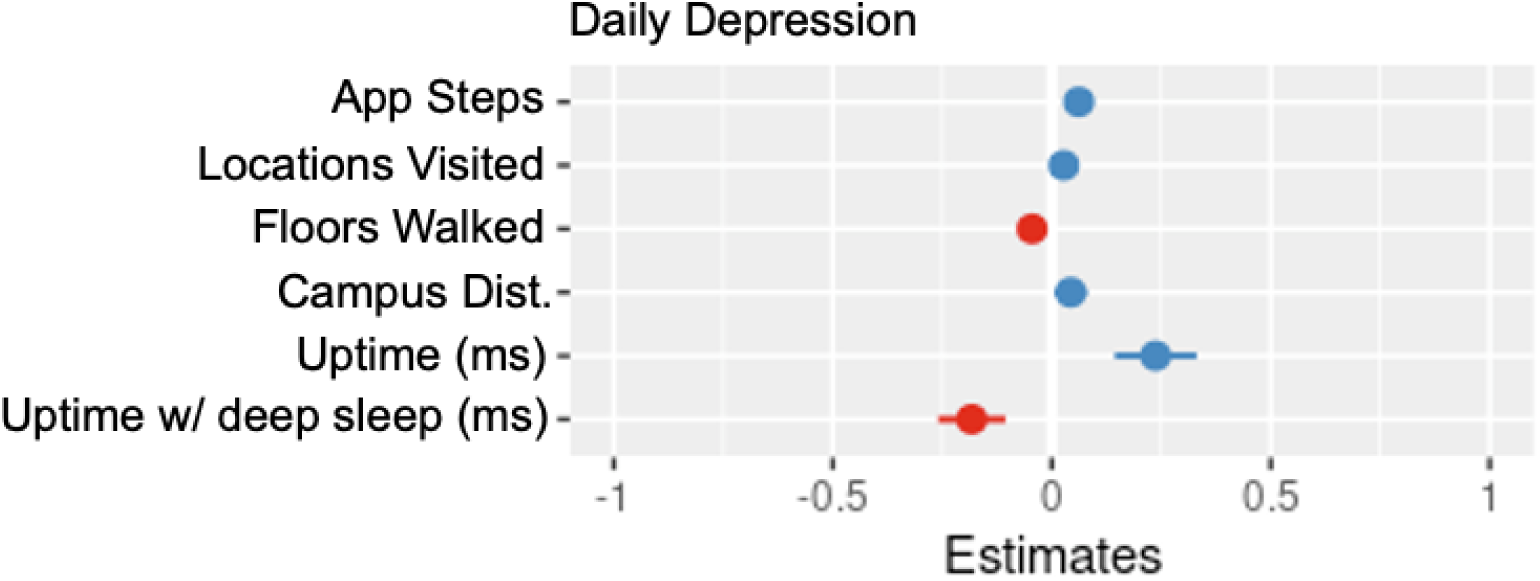
Coefficient plot for parameters of Model #5 – daily depression.

App steps and locations visited had minor positive associations with daily depression. There was a positive association with system uptime and daily depression, and a negative association with system sleep time and depression.

**Figure 10.**
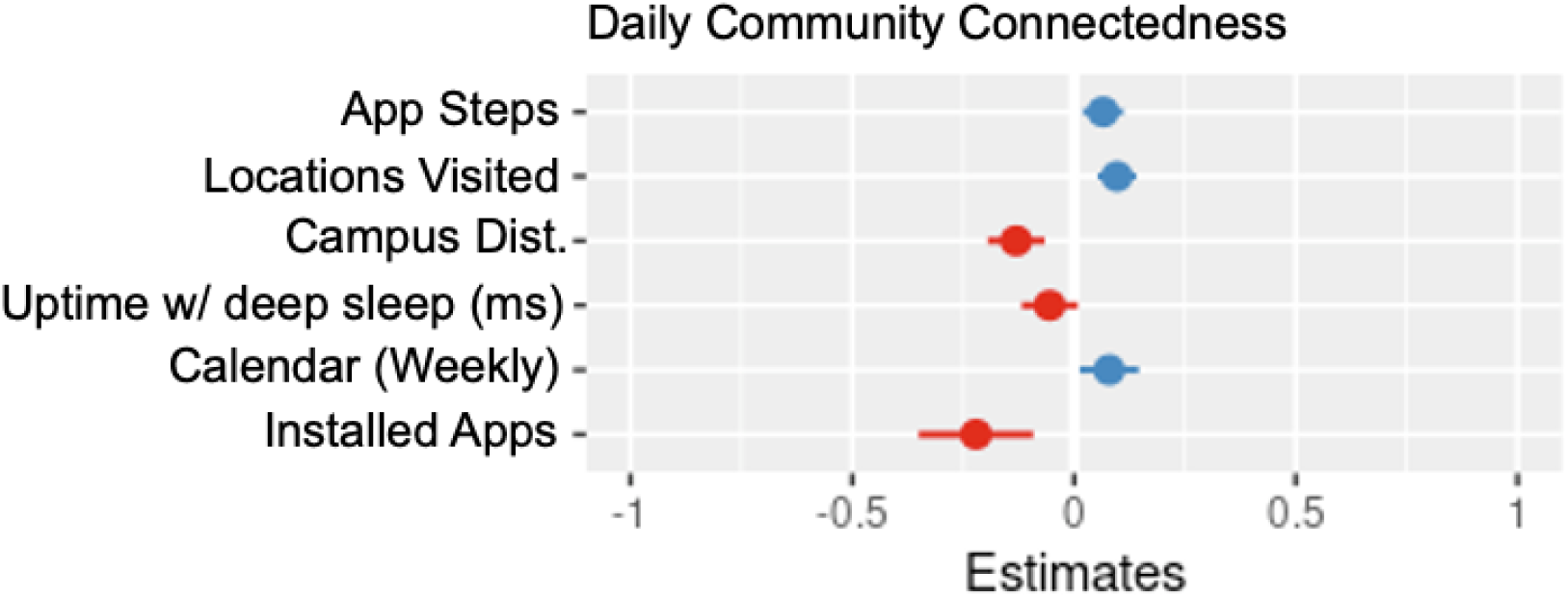
Coefficient plot for parameters of Model #6 – daily community connectedness.

App steps, locations visited, and proximity to campus had positive associations with daily community connectedness. There was a negative association with the number of installed apps and system sleep time with community connectedness. The number of weekly calendar events also had a positive association with community connectedness.

#### Regressing daily questions 7-9 on device digital measures

Table 8 shows the results of three mixed linear models relating device digital measures to a second daily measure of anxiety, a daily measure of distress, and a second daily measure of resilience.

**Table 8.** First round of mixed linear models for daily anxiety, distress, and resilience.

|  | Dependent variable |  |  |  |  |  |
| --- | --- | --- | --- | --- | --- | --- |
|  | Daily Q7 |  | Daily Q8 |  | Daily Q9 |  |
| Measure | Anxiety #2 |  | Distress |  | Resilience #2 |  |
| Model # | 7 |  | 8 |  | 9 |  |
| Observations | 2494 |  | 2494 |  | 2494 |  |
| Participants | 46 |  | 46 |  | 46 |  |
| Device digital measure | Parameter (SE) | P-value | Parameter (SE) | P-value | Parameter (SE) | P-value |
| Step count (weekly) | 0.099 (0.031) | 0.001 | -0.266 (0.049) | 7.11E-08 | -0.1 (0.026) | 0.000 |
| GPS locations visited | 0.087 (0.024) | 0.000 | -0.134<br>(0.038) | 0.000 | 0.122 (0.021) | 3.7E-09 |
| Floors walked (up + down) | 0.057 (0.028) | 0.039 | 0.089<br>(0.044) | 0.041 | 0.107 (0.023) | 0.000 |
| Campus Dist. (less is closer to UWO) | -0.085<br>(0.036) | 0.022 | -0.201<br>(0.057) | 0.000 | -0.131<br>(0.031) | 0.000 |
| System uptime (ms) | 0.027 (0.099) | 0.784 | 0.111<br>(0.154) | 0.470 | 0.19 (0.083) | 0.023 |
| System uptime with deep sleep (ms) | 0.113 (0.077) | 0.146 | -0.133<br>(0.12) | 0.267 | -0.167<br>(0.065) | 0.01 |
| Available RAM (bytes) | 0.012 (0.06) | 0.84 | -0.022<br>(0.094) | 0.815 | -0.01 (0.05) | 0.838 |
| Free space (percent) | 0.059 (0.053) | 0.271 | 0.031<br>(0.08) | 0.693 | -0.091<br>(0.045) | 0.045 |
| Time spent using SHC 2.0 app (ms) | -0.056<br>(0.022) | 0.012 | 0.064<br>(0.035) | 0.067 | -0.05 (0.018) | 0.006 |
| Calendar count (daily) | 0.028 (0.038) | 0.462 | -0.243<br>(0.06) | 5.85E-05 | -0.05 (0.033) | 0.124 |
| Calendar<br>count<br>(weekly) | -0.17 (0.042) | 0.000 | 0.207<br>(0.07) | 0.001 | 0.023 (0.036) | 0.52 |
| Installed apps<br>(total /<br>List of 40) | 0.192 (0.07) | 0.006 | 0.07<br>(0.105) | 0.502 | 0.17 (0.06) | 0.004 |

#### Revised Models #7-9 using significant parameters only, with coefficient plots

**Table 9.** Revised second round (using only significant digital measure items) of mixed linear models for anxiety measure #2, distress, and resilience measure #2.

|  | Dependent variable |  |  |
| --- | --- | --- | --- |
|  | Anxiety #2 | Distress | Resilience #2 |
| Revised Model # | 7 | 8 | 9 |
| Observations | 2514 | 2514 | 2494 |
| Participants | 46 | 46 | 46 |
| Device digital measure | Parameter (SE) | Parameter (SE) | Parameter (SE) |
| Step count<br>(weekly) | 0.08 (0.030) | -0.27 (0.048) | -0.099 (0.026) |
| GPS locations visited | 0.110 (0.023) | -0.142 (0.036) | 0.12 (0.02) |
| Floors walked (up + down) | 0.07 (0.027) | 0.091 (0.043) | 0.105 (0.023) |
| Campus Dist. (less is closer to UWO) | -0.098 (0.035) | -0.203 (0.055) | -0.13 (0.031) |
| System uptime (ms) |  |  | 0.19 (0.083) |
| System uptime with deep sleep (ms) |  |  | -0.165 (0.065) |
| Free space (percent) |  |  | -0.091 (0.045) |
| Time spent using SHC 2.0 app (ms) | -0.048 (0.02) |  | -0.051 (0.018) |
| Calendar count (daily) |  | -0.24 (0.060) |  |
| Calendar count (weekly) | -0.155 (0.036) | 0.21 (0.065) |  |
| Installed apps (total / List of 40) | 0.195 (0.0695) |  | 0.169 (0.059) |

**Figure 11.**
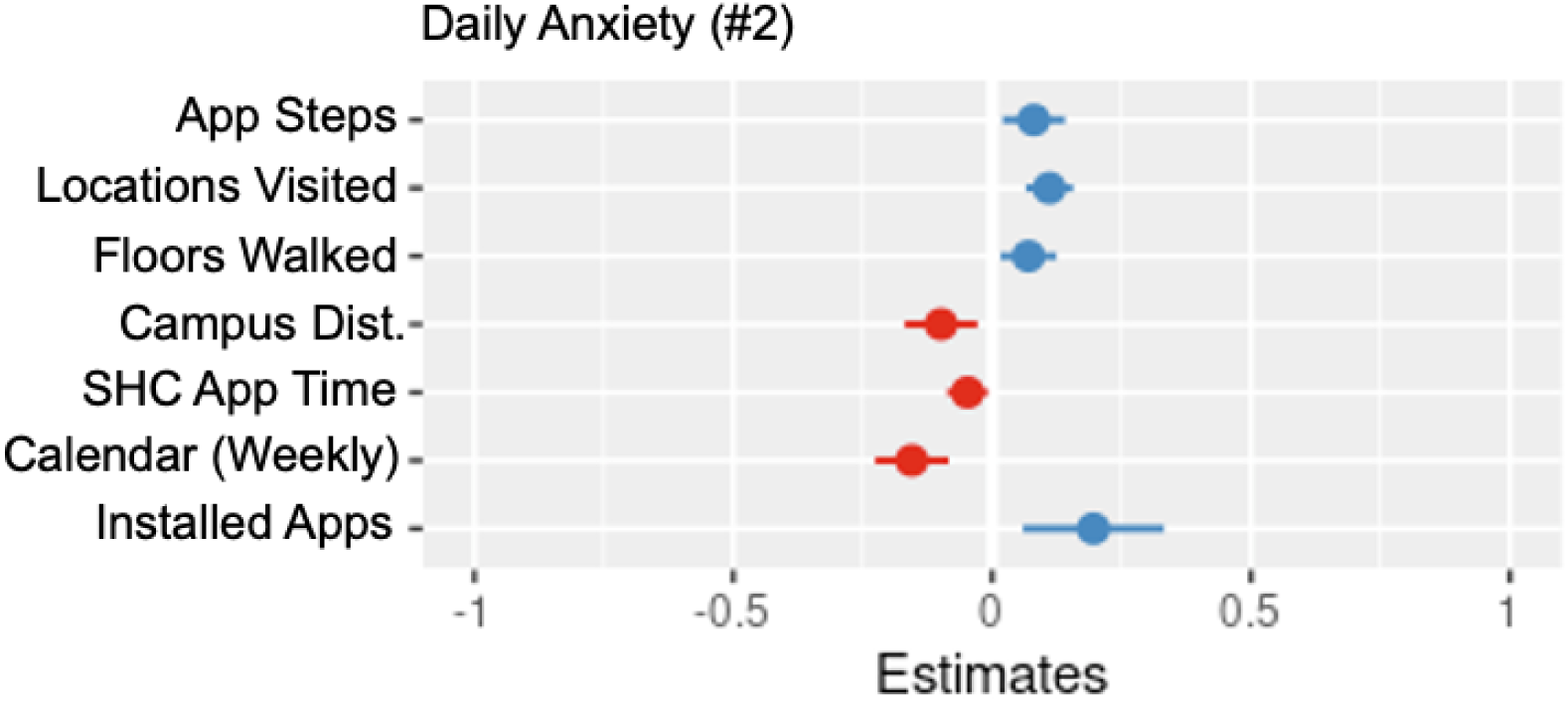
Coefficient plot for parameters of Model #7 – daily anxiety measure #2.

Physical activity (app steps, locations visited, floors walked) and proximity to campus had slight positive associations with anxiety, while installed apps had a more positive association with anxiety. There was a negative association with weekly calendar events.

**Figure 12.**
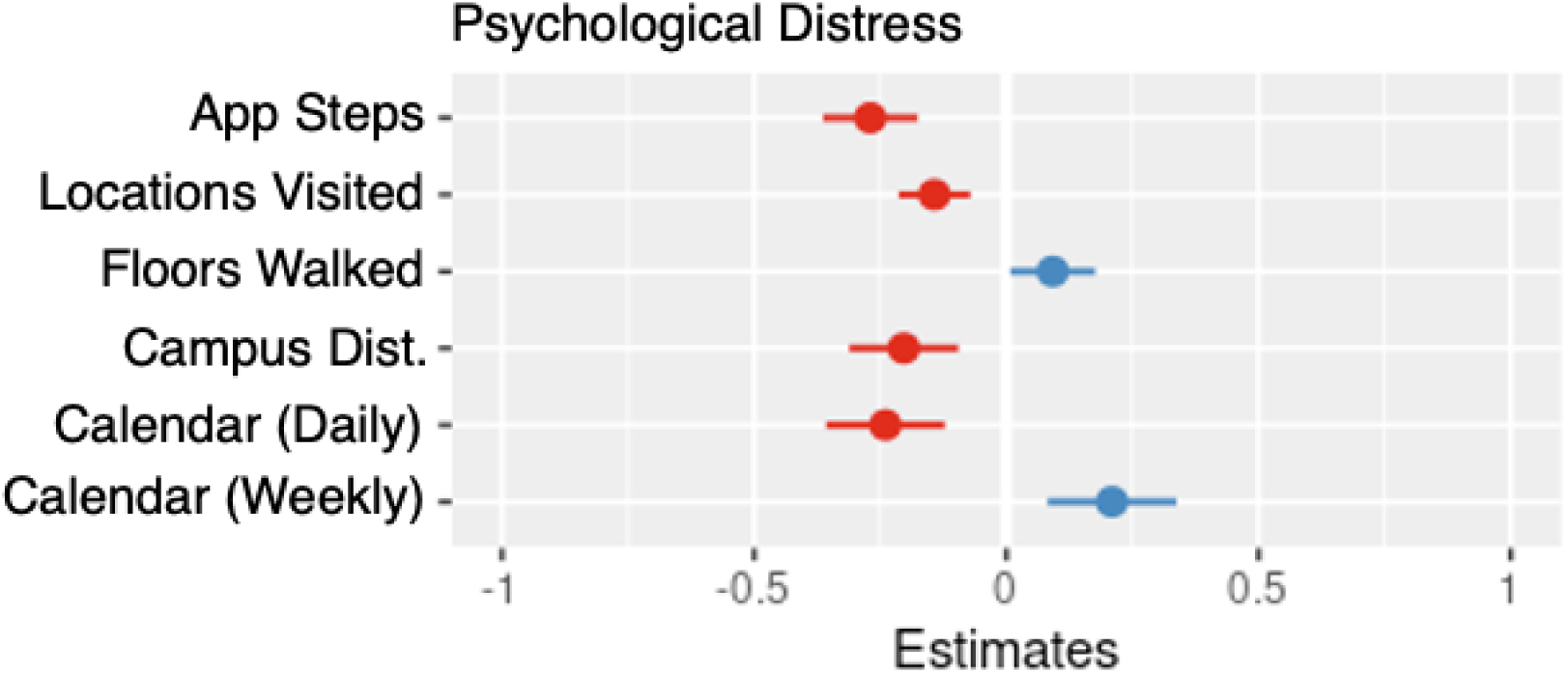
Coefficient plot for parameters of Model #8 – Psychological Distress

Some measures of physical activity (app steps, locations visited) and daily calendar events had a negative association with distress. Proximity to campus, floors walked, and weekly calendar events had a positive association with distress.

**Figure 13.**
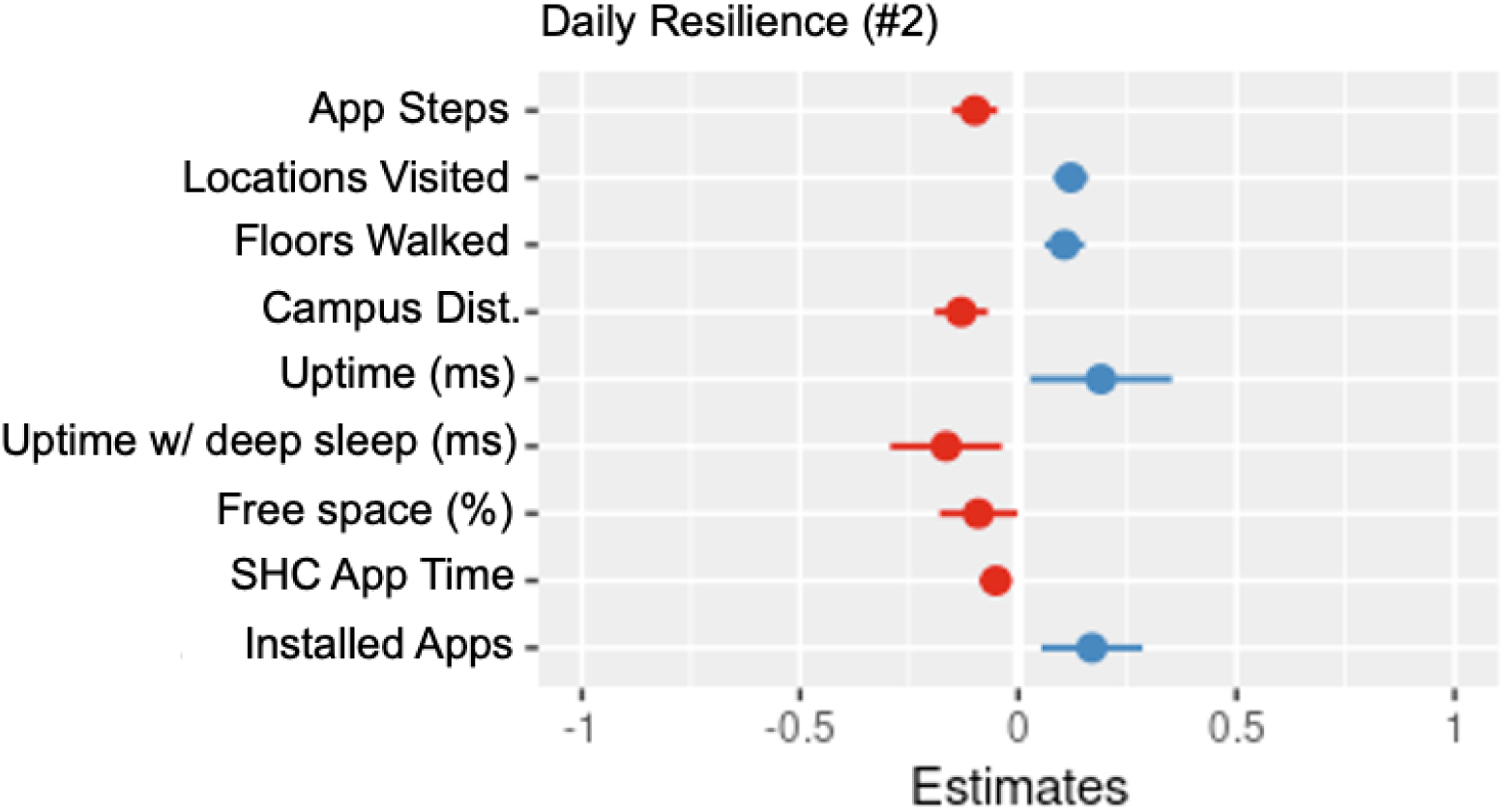
Coefficient plot for parameters of Model #9 – daily resilience measure #2.

App steps had a slight negative association with the second daily measure of resilience. System sleep time, free space also had negative associations with this measure of daily resilience. Locations visited, floors walked, proximity to campus, system uptime, and installed app count all had positive associations with this measure of resilience.

#### Result of Model #10: Regressing daily total questionnaire sum on device digital measures

**Table 10.** Revised second round (using only significant device digital measure items) of mixed linear model relating daily questionnaire sum to device digital measures. The first model is omitted for brevity. The initial model is included as an Appendix.

|  |  |
| --- | --- |
|  | Dependent variable |
|  | Daily questionnaire variant sum |
| Revised Model # | 10 |
| Observations | 2514 |
| Participants | 46 |
| Device digital measure | Parameter (SE) |
| Floors walked (up + down) | 0.669 (0.118) |
| Location flag | -0.576 (0.186) |
| System uptime (ms) | -0.56 (0.226) |
| Calendar count (daily) | -0.47 (0.18) |
| Installed apps (total / List of 40) | 0.72 (0.38) |

**Figure 14.**
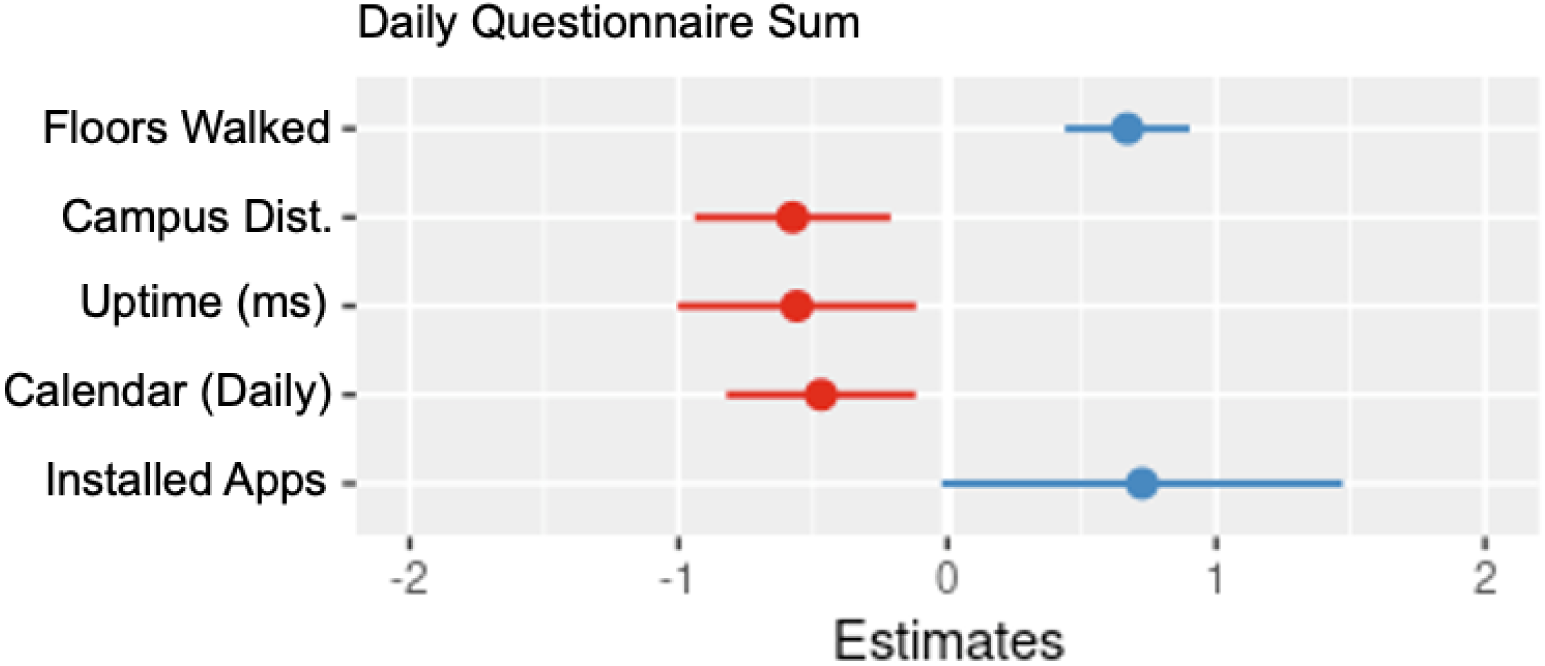
Coefficient plot for parameters of Model #10 – daily questionnaire sum.

Floors walked, proximity to campus, and installed app count had positive associations with the daily questionnaire sum. System uptime and daily calendar events had negative associations with the daily questionnaire sum.

### Mixed linear models of SHC 2.0 weekly questionnaire variant

In this section, the mixed linear model fits for the measures of the SHC 2.0 weekly questionnaire variant are presented. Note that for several weekly questions, mixed linear model fits did not yield any significant associations. This was the case for questions: 2 (psychological well-being), 5 (depression), and 6 (community connectedness).

#### Regressing weekly questions 1, 3, and 4 on device digital measures

Table 11 shows the results of three mixed linear models relating device digital measures to weekly life satisfaction, a first weekly measure of resilience, and a first weekly measure of anxiety.

**Table 11.** First round of mixed linear models for weekly life satisfaction, weekly resilience, and weekly anxiety.

|  | Dependent variable |  |  |  |  |  |
| --- | --- | --- | --- | --- | --- | --- |
|  | Weekly Q1 |  | Weekly Q3 |  | Weekly Q4 |  |
|  | Life Satisfaction |  | Resilience #1 |  | Anxiety #1 |  |
| Model # | 11 |  | 12 |  | 13 |  |
| Observations | 992 |  | 992 |  | 1003 |  |
| Participants | 44 |  | 44 |  | 44 |  |
| Device digital measure | Parameter (SE) | P-value | Parameter (SE) | P-value | Parameter (SE) | P-value |
| Step count (weekly) | 0.107 (0.058) | 0.066 | 0.0683 (0.041) | 0.096 | 0.022 (0.032) | 0.498 |
| GPS locations visited | 0.014 (0.043) | 0.747 | -0.098 (0.03) | 0.001 | 0.077 (0.023) | 0.001 |
| Floors walked<br>(up + down) | 0.032 (0.065) | 0.620 | -0.113<br>(0.046) | 0.013 | -0.182<br>(0.035) | 3.31E-07 |
| Campus Dist.<br>(less is closer<br>to UWO) | -0.258 (0.07) | 0.000 | 0.184<br>(0.049) | 0.000 | 0.075 (0.04) | 0.048 |
| System<br>uptime (ms) | -0.696<br>(0.245) | 0.004 | 0.193<br>(0.172) | 0.259 | 0.022 (0.133) | 0.865 |
| System<br>uptime with<br>deep sleep<br>(ms) | 0.461 (0.165) | 0.005 | -0.044<br>(0.116) | 0.707 | -0.073<br>(0.089) | 0.412 |
| Available<br>RAM (bytes) | 0.072 (0.102) | 0.478 | 0.064<br>(0.072) | 0.37 | -0.062<br>(0.055) | 0.260 |
| Free space<br>(percent) | -0.173<br>(0.168) | 0.304 | 0.303<br>(0.1171) | 0.01 | 0.061 (0.085) | 0.471 |
| Time spent<br>using SHC 2.0<br>app (ms) | -0.036<br>(0.067) | 0.589 | -0.014<br>(0.047) | 0.763 | 0.059 (0.036) | 0.108 |
| Calendar<br>count (daily) | 0.164 (0.064) | 0.011 | 0.012<br>(0.045) | 0.786 | -0.039<br>(0.035) | 0.269 |
| Calendar<br>count<br>(weekly) | -0.152<br>(0.109) | 0.163 | 0.008<br>(0.076) | 0.916 | -0.093<br>(0.059) | 0.115 |
| Installed apps<br>(total /<br>List of 40) | 0.766 (0.155) | 0.000 | -0.369<br>(0.108) | 0.000 | -0.182 (0.08) | 0.025 |

#### Revised Models #11-13 using significant parameters only, with coefficient plots

**Table 12.**
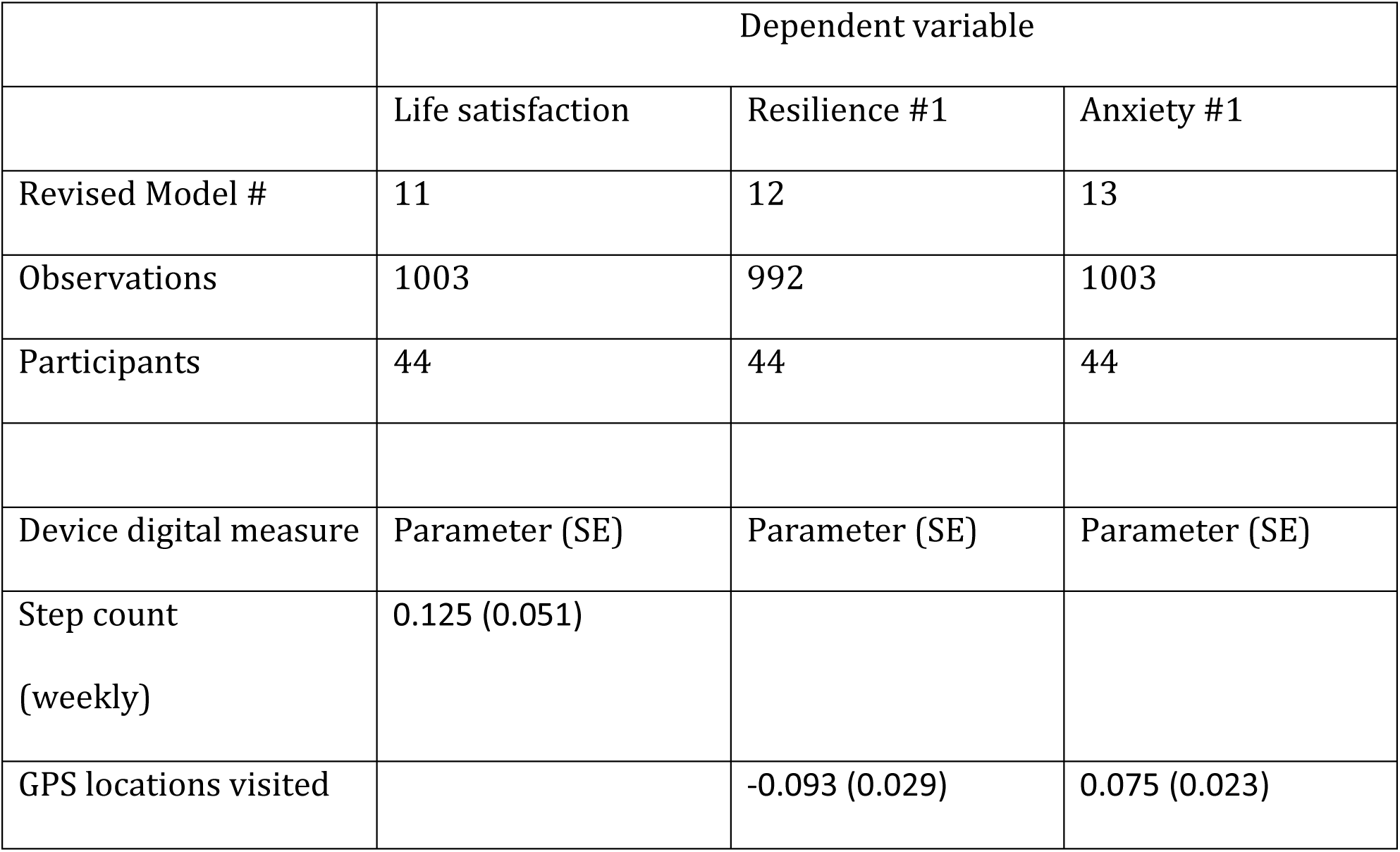
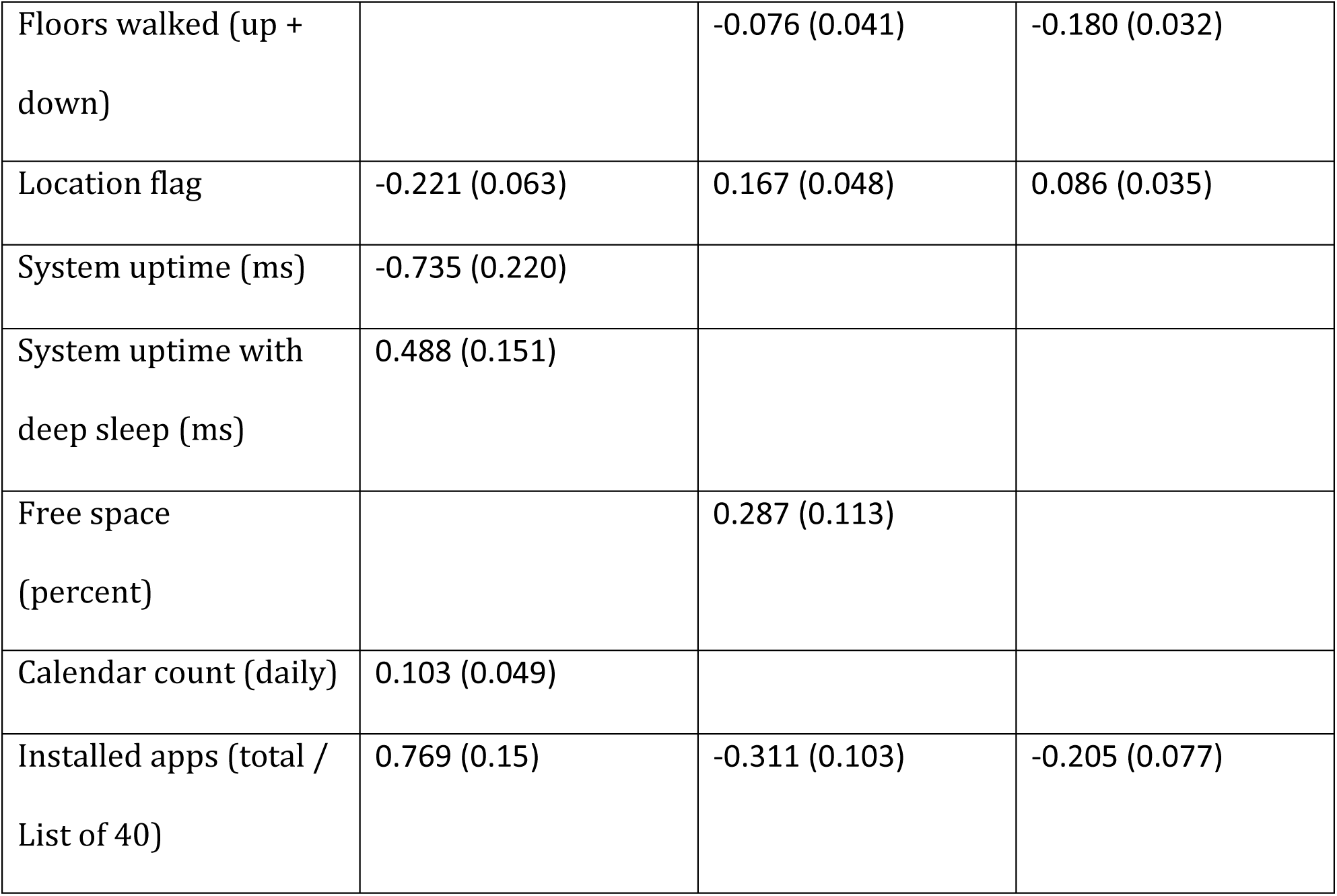
Revised second round (using only significant digital measure items) of mixed linear models for weekly life satisfaction, weekly resilience measure #1, weekly anxiety measure #1.

**Figure 15.**
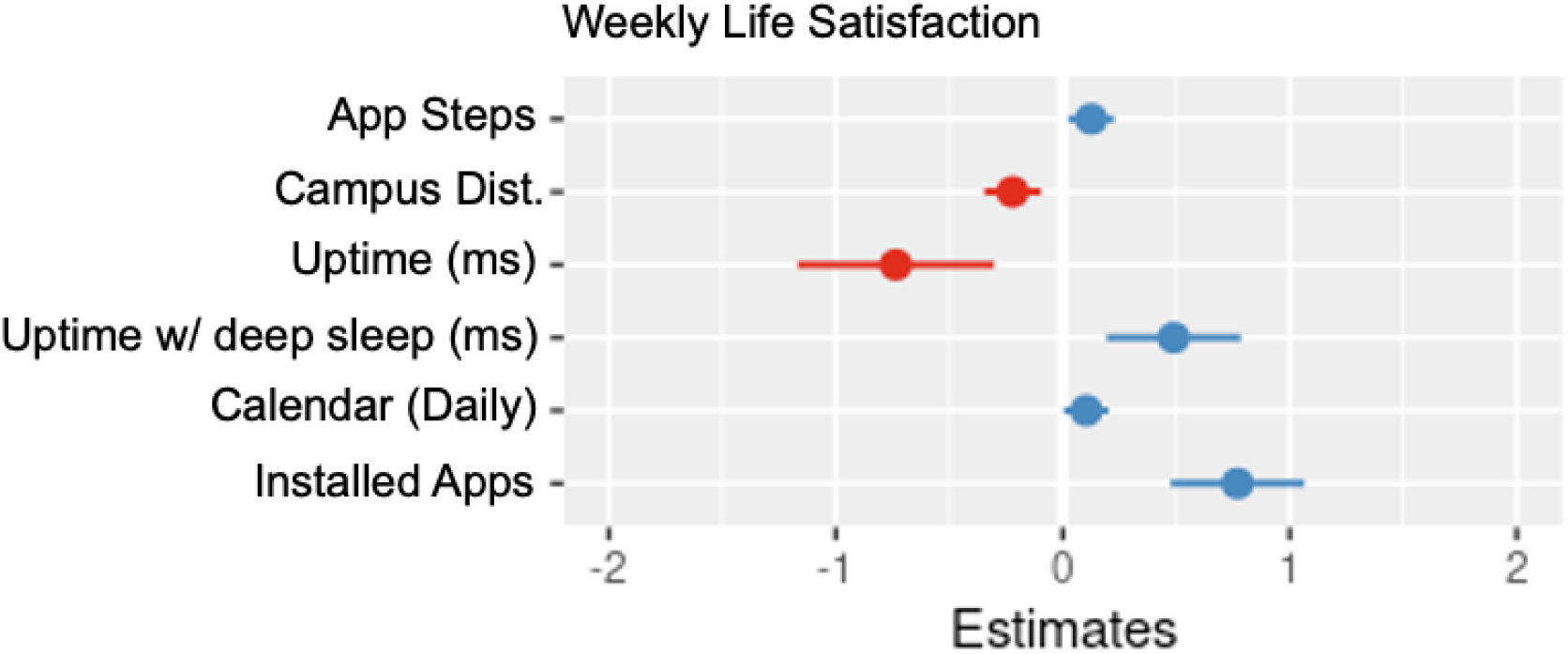
Coefficient plot for parameters of Model #11 – weekly life satisfaction.

App steps, calendar events on the day the weekly variant was completed, proximity to campus, and installed app count all had positive associations with weekly life satisfaction. There was a positive association with system sleep time which may be indicative of participants getting more sleep. There was a corresponding negative association with system uptime.

**Figure 16.**
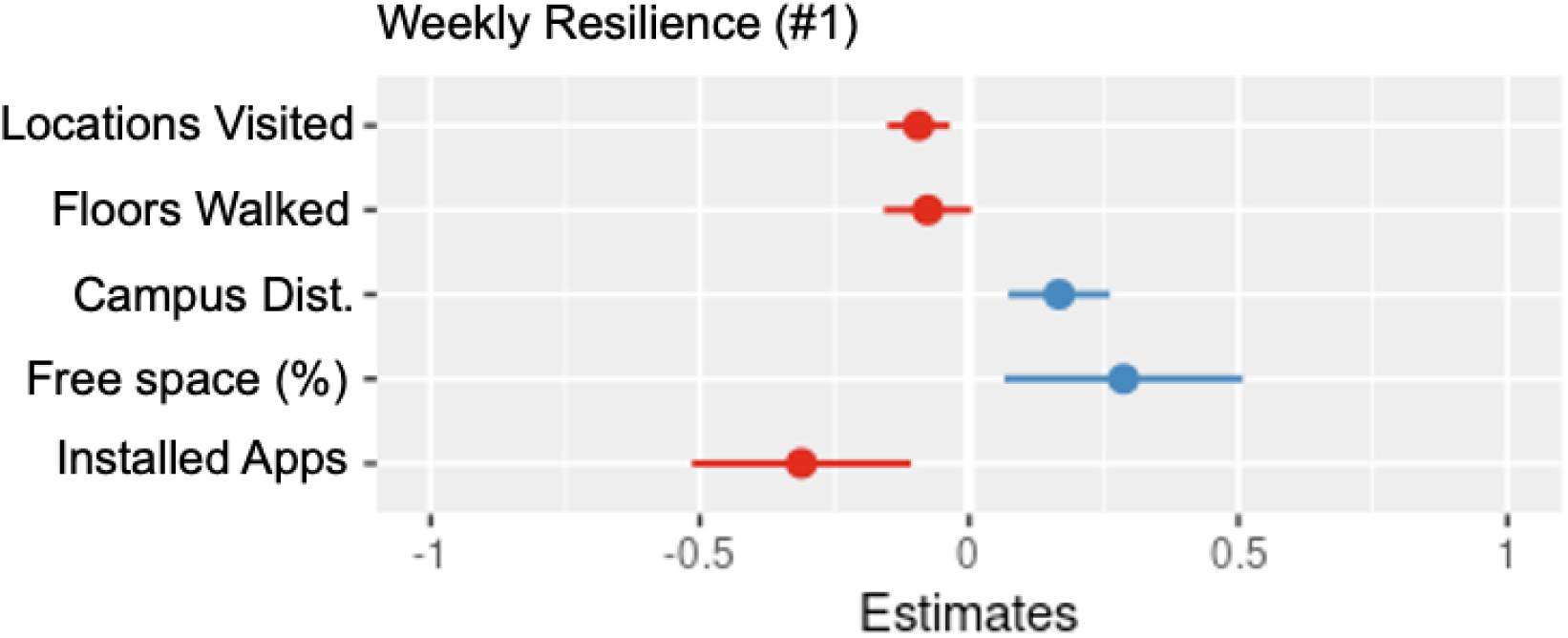
Coefficient plot for parameters of Model #12 – weekly resilience measure #1.

The first measure of weekly resilience had a positive association with distance from campus and device free space. Weekly resilience had a negative association with installed app count, locations visited, and floors walked.

**Figure 17.**
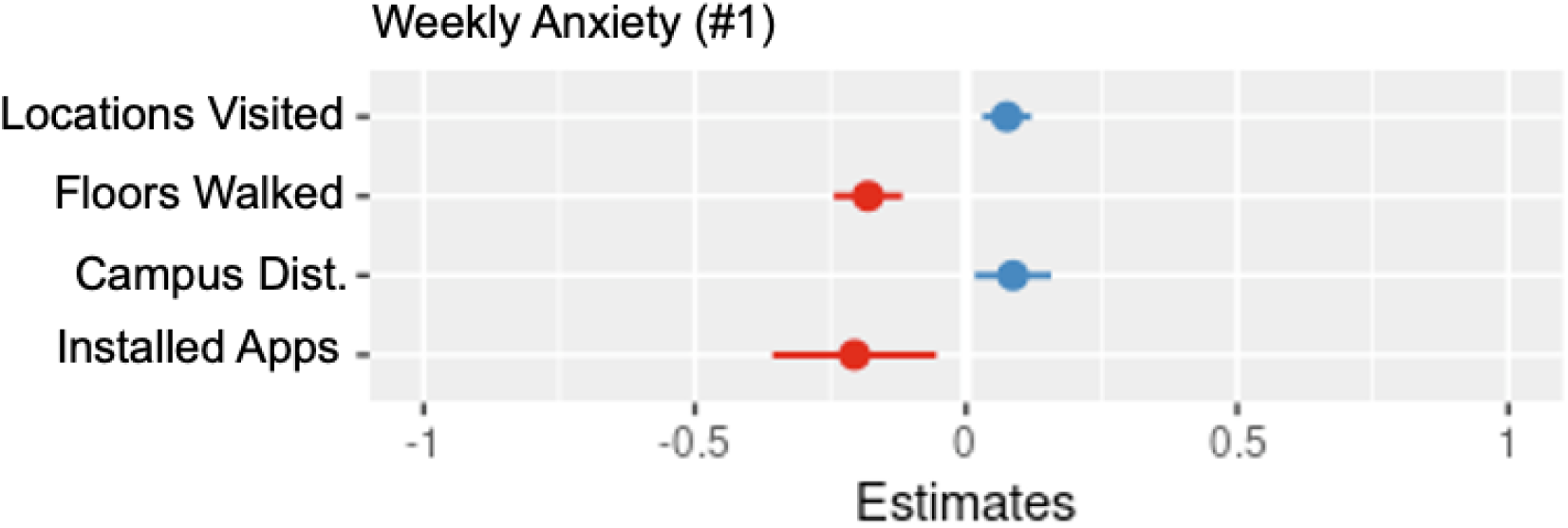
Coefficient plot for parameters of Model #13 – weekly anxiety measure #1.

There was a slight positive association with locations visited and weekly anxiety. This weekly anxiety measure had negative associations with floors walked, installed app count, and proximity to campus.

#### Regressing weekly questions 7, 8, and 9 on device digital measures

Table 13 shows the results of three mixed linear models relating device digital measures to weekly physical activity, a second measure of anxiety, and a second measure of resilience.

**Table 13.**
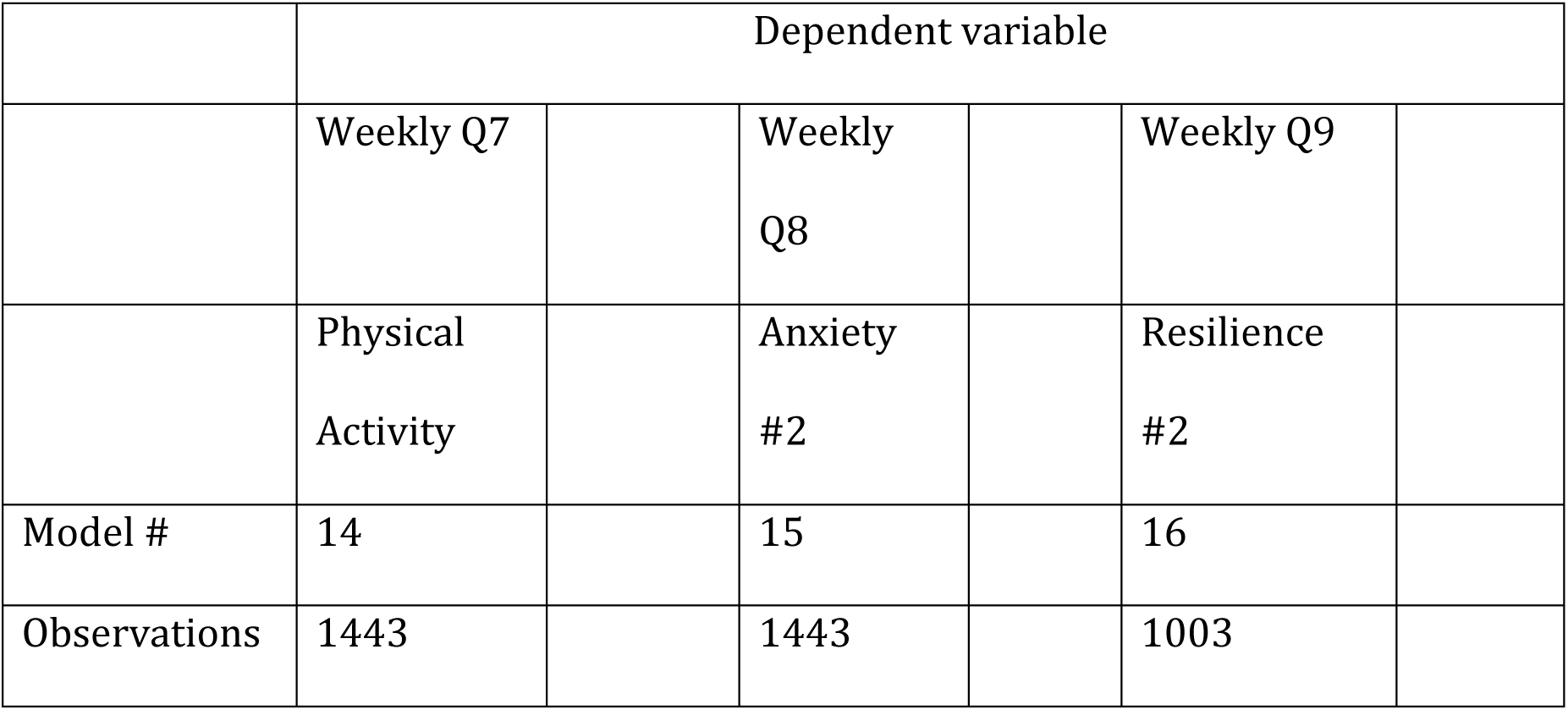
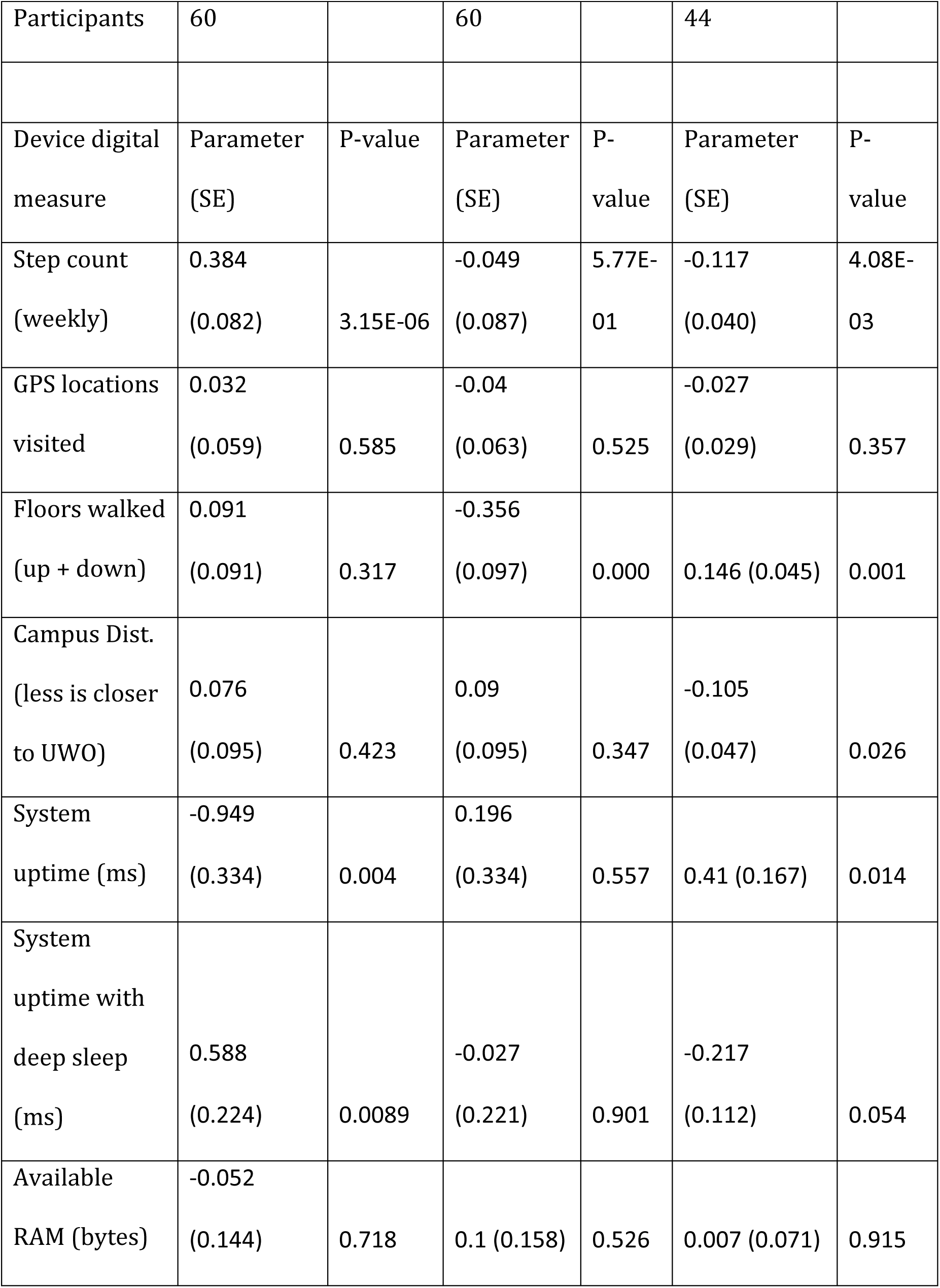
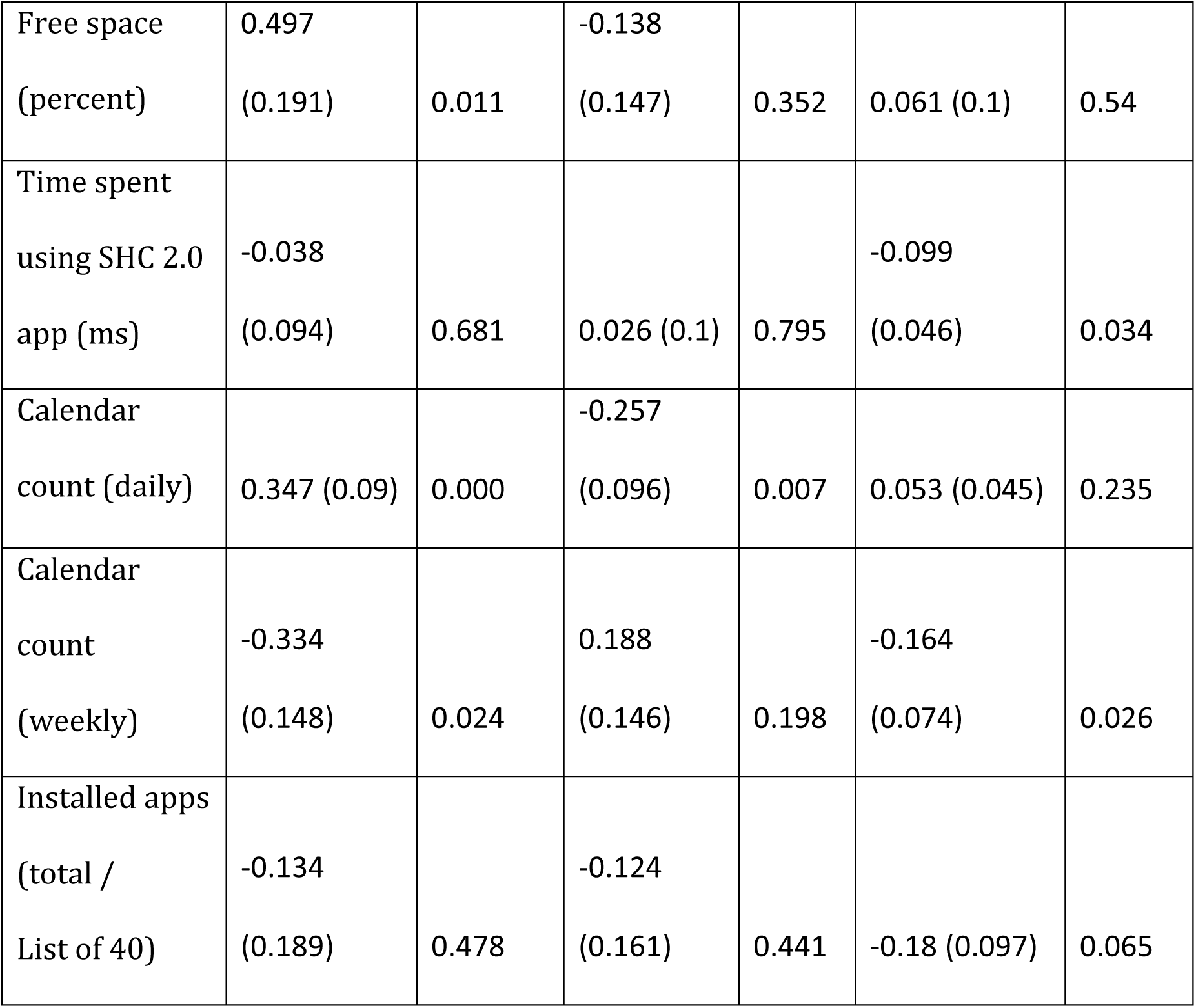
First round of mixed linear models for weekly physical activity, weekly anxiety, and weekly resilience.

|  | Dependent variable |  |  |  |  |
| --- | --- | --- | --- | --- | --- |
|  | Weekly Q7 |  | Weekly Q8 |  | Weekly Q9 |
|  | Physical Activity |  | Anxiety #2 |  | Resilience #2 |
| Model # | 14 |  | 15 |  | 16 |
| Observations | 1443 |  | 1443 |  | 1003 |

|  |  |  |  |  |  |  |
| --- | --- | --- | --- | --- | --- | --- |
| Participants | 60 |  | 60 |  | 44 |  |
| Device digital measure | Parameter (SE) | P-value | Parameter (SE) | P-value | Parameter (SE) | P-value |
| Step count (weekly) | 0.384 (0.082) | 3.15E-06 | -0.049 (0.087) | 5.77E-01 | -0.117 (0.040) | 4.08E-03 |
| GPS locations visited | 0.032 (0.059) | 0.585 | -0.04 (0.063) | 0.525 | -0.027 (0.029) | 0.357 |
| Floors walked (up + down) | 0.091 (0.091) | 0.317 | -0.356 (0.097) | 0.000 | 0.146 (0.045) | 0.001 |
| Campus Dist. (less is closer to UW0) | 0.076 (0.095) | 0.423 | 0.09 (0.095) | 0.347 | -0.105 (0.047) | 0.026 |
| System uptime (ms) | -0.949 (0.334) | 0.004 | 0.196 (0.334) | 0.557 | 0.41 (0.167) | 0.014 |
| System uptime with deep sleep (ms) | 0.588 (0.224) | 0.0089 | -0.027 (0.221) | 0.901 | -0.217 (0.112) | 0.054 |
| Available RAM (bytes) | -0.052 (0.144) | 0.718 | 0.1 (0.158) | 0.526 | 0.007 (0.071) | 0.915 |

|  |  |  |  |  |  |  |
| --- | --- | --- | --- | --- | --- | --- |
| Free space<br>(percent) | 0.497<br>(0.191) | 0.011 | -0.138<br>(0.147) | 0.352 | 0.061 (0.1) | 0.54 |
| Time spent<br>using SHC 2.0<br>app (ms) | -0.038<br>(0.094) | 0.681 | 0.026 (0.1) | 0.795 | -0.099<br>(0.046) | 0.034 |
| Calendar<br>count (daily) | 0.347 (0.09) | 0.000 | -0.257<br>(0.096) | 0.007 | 0.053 (0.045) | 0.235 |
| Calendar<br>count<br>(weekly) | -0.334<br>(0.148) | 0.024 | 0.188<br>(0.146) | 0.198 | -0.164<br>(0.074) | 0.026 |
| Installed apps<br>(total /<br>List of 40) | -0.134<br>(0.189) | 0.478 | -0.124<br>(0.161) | 0.441 | -0.18 (0.097) | 0.065 |

#### Revised Models #14-16 using significant parameters only, with coefficient plots

**Table 14.**
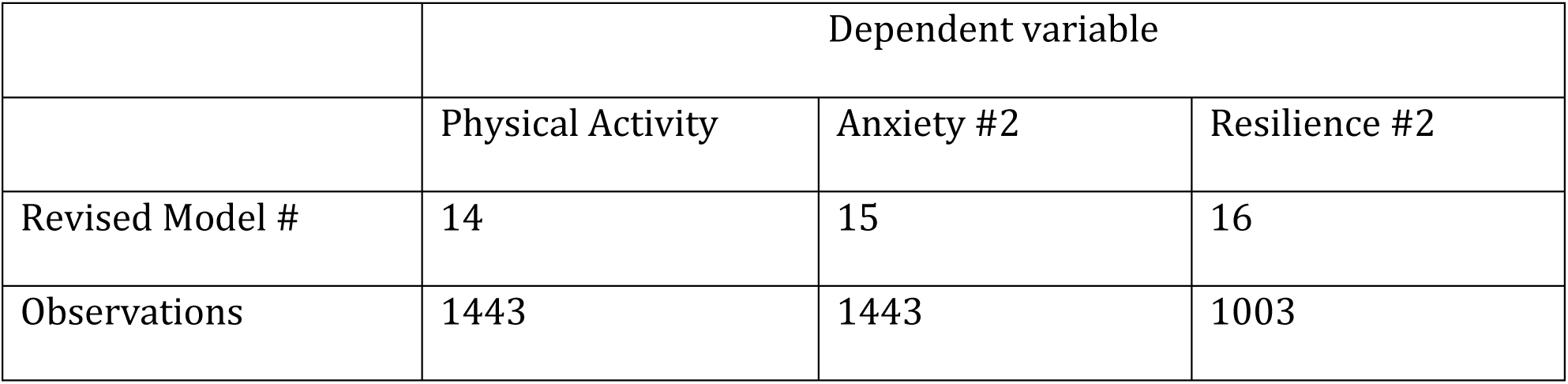
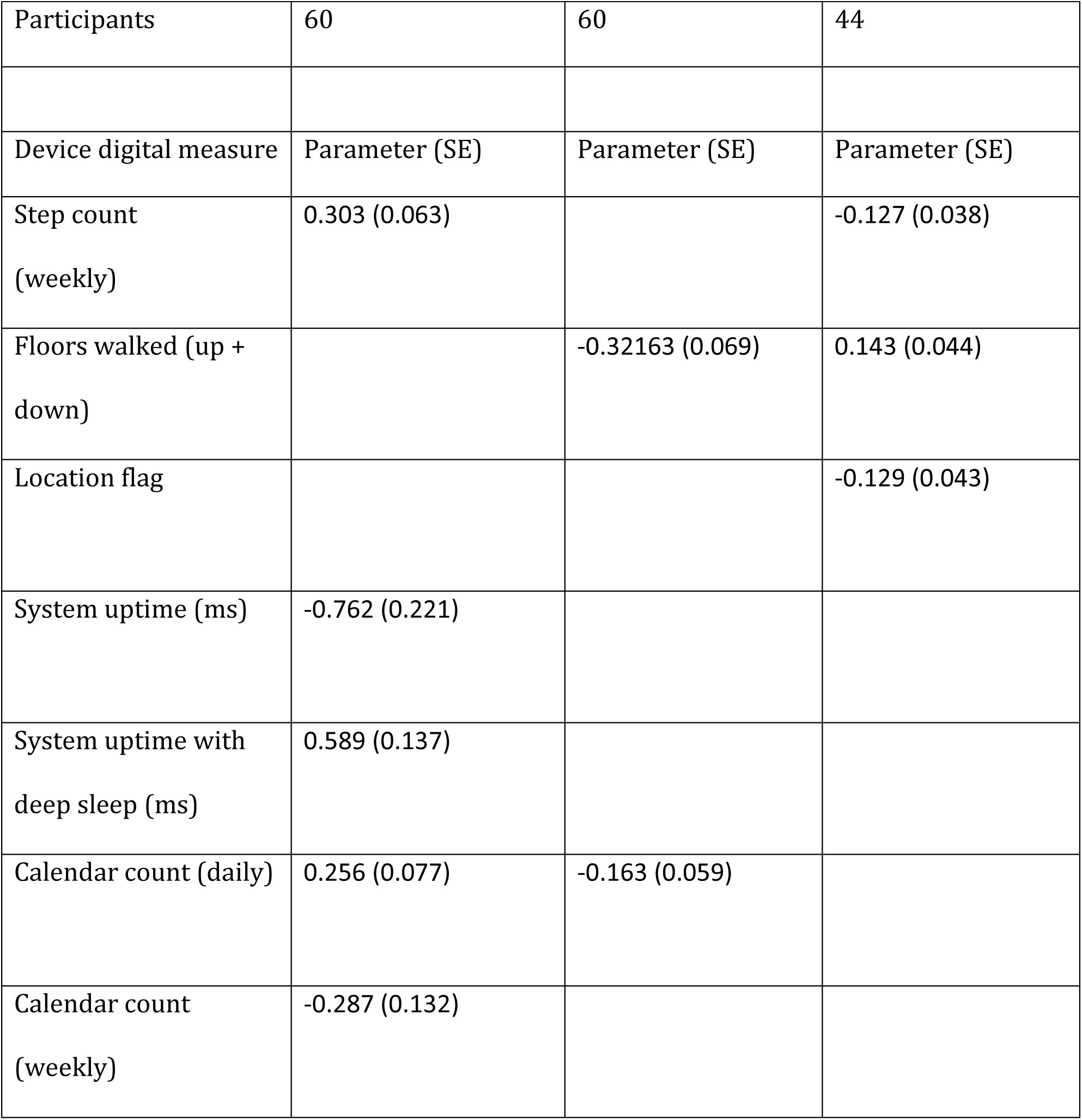
Revised second round (using only significant digital measure items) of mixed linear models for weekly physical activity, a second weekly anxiety measure, and a second weekly measure of resilience.

|  | Dependent variable |  |  |
| --- | --- | --- | --- |
|  | Physical Activity | Anxiety #2 | Resilience #2 |
| Revised Model # | 14 | 15 | 16 |
| Observations | 1443 | 1443 | 1003 |
| Participants | 60 | 60 | 44 |
| Device digital measure | Parameter (SE) | Parameter (SE) | Parameter (SE) |
| Step count<br>(weekly) | 0.303 (0.063) |  | -0.127 (0.038) |
| Floors walked (up +<br>down) |  | -0.32163 (0.069) | 0.143 (0.044) |
| Location flag |  |  | -0.129 (0.043) |
| System uptime (ms) | -0.762 (0.221) |  |  |
| System uptime with<br>deep sleep (ms) | 0.589 (0.137) |  |  |
| Calendar count (daily) | 0.256 (0.077) | -0.163 (0.059) |  |
| Calendar count<br>(weekly) | -0.287 (0.132) |  |  |

**Figure 18.**
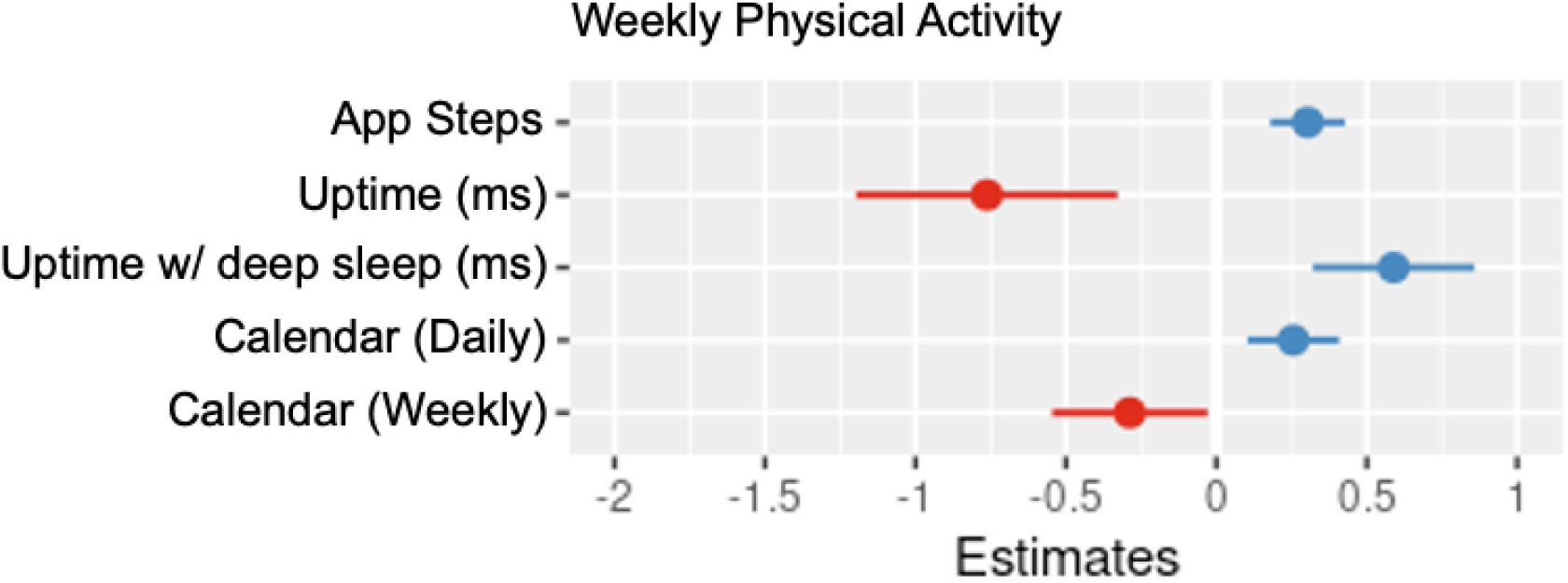
Coefficient plot for parameters of Model #14 – weekly physical activity.

As expected, app steps had a positive association with weekly physical activity. There was also a significant negative association in system up time, but a positive association with system sleep time, which may be indicative of physically active participants getting more sleep. Physically active participants had associations with more events on the day they completed the weekly survey, but a negative association with events throughout their week.

**Figure 19.**
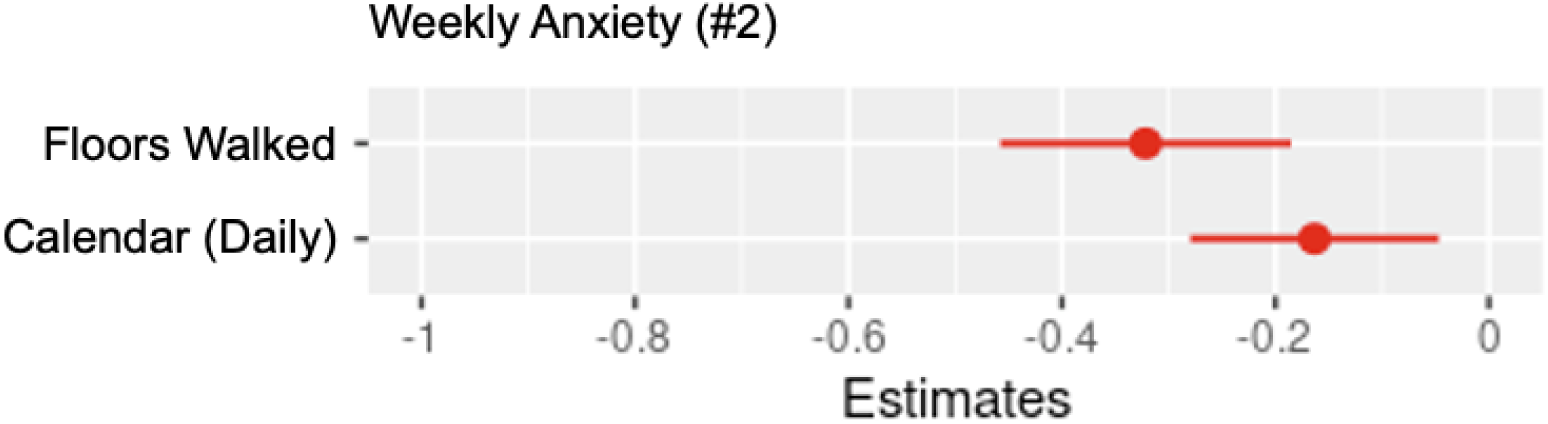
Coefficient plot for parameters of Model #15 – weekly anxiety measure #2.

Like the first weekly anxiety measure, this measure of anxiety had a negative association with floors walked. Calendar events, on the day the weekly questionnaire was answered, also had a negative association with anxiety in this second measure.

**Figure 20.**
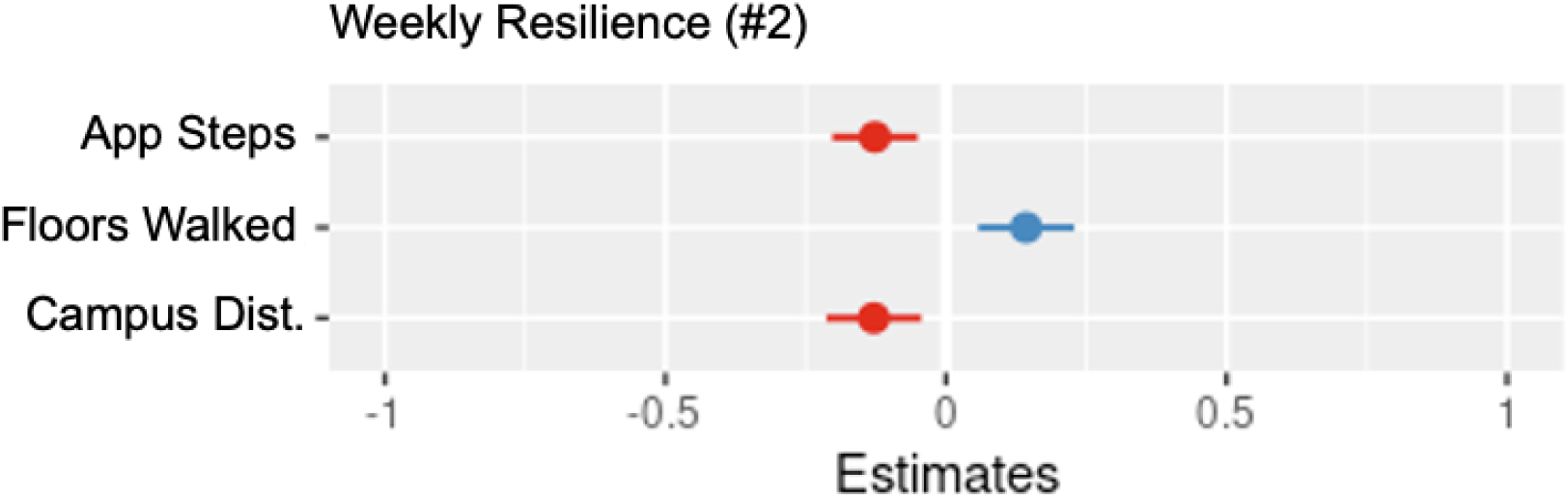
Coefficient plot for parameters of Model #16 – weekly resilience measure #2.

App steps had a negative association with this second measure of weekly resilience. There was a positive association with floors walked, and proximity to campus.

#### Result of Model #17: Regressing weekly total questionnaire sum on device digital measures

**Table 15.** Revised second round (using only significant digital measure items) of mixed linear model for weekly questionnaire sum and device digital measures. The first model is omitted for brevity. The initial model is included as an Appendix.

|  |  |
| --- | --- |
|  | Dependent variable |
|  | Weekly<br>questionnaire<br>variant sum |
| Revised Model # | 17 |
| Observations | 1003 |
| Participants | 44 |
| Device digital measure | Parameter (SE) |
| Floors walked (up +<br>down) | 0.748 (0.207) |
| Campus Dist. (less is<br>closer to UW0) | -0.881 (0.230) |
| System uptime (ms) | -2.789 (0.802) |
| System uptime with<br>deep sleep (ms) | 1.730 (0.545) |
| Calendar count (daily) | 0.492 (0.180) |
| Installed apps (total /<br>List of 40) | 1.644 (0.553) |

**Figure 21.**
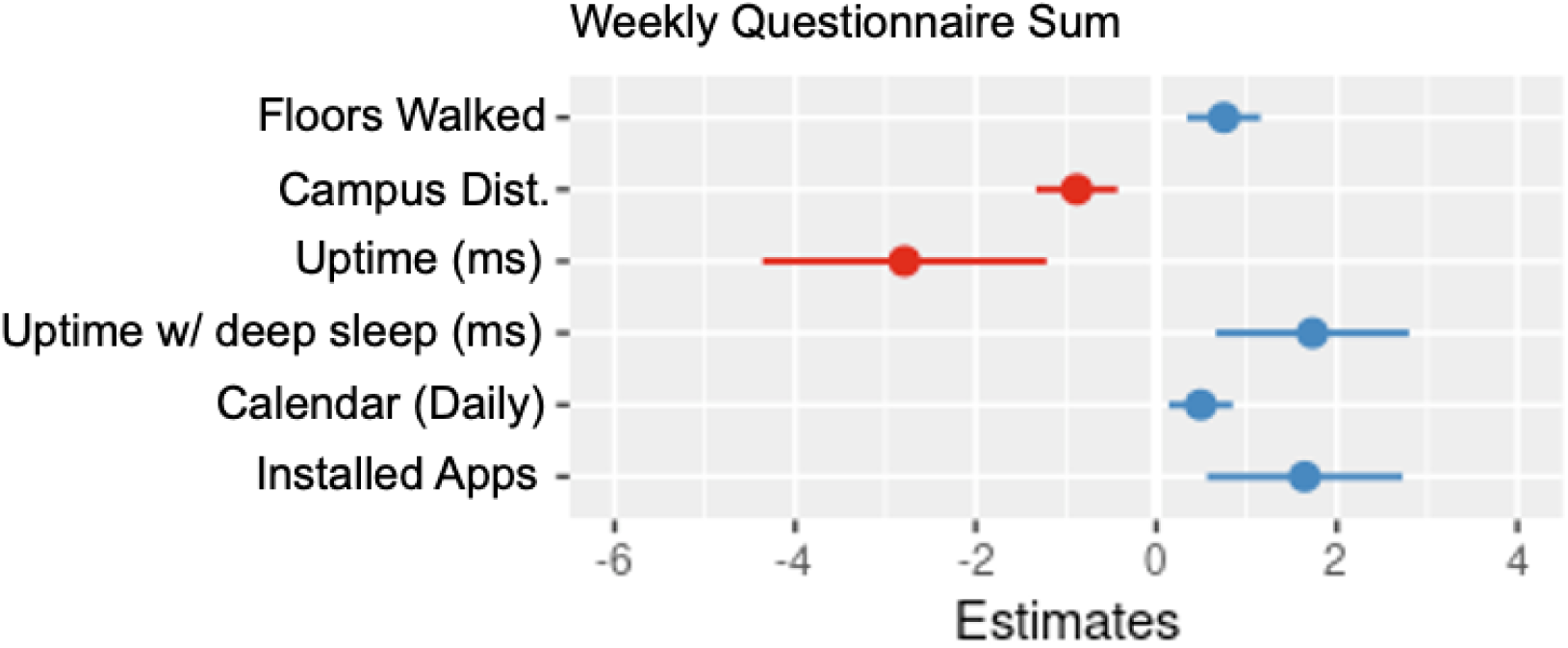
Coefficient plot for parameters of Model #17 – sum of weekly questionnaire.

Floors walked, proximity to campus, system sleep time, installed apps, and to a lesser extent, calendar events the day this was answered, all had positive associations with the weekly questionnaire sum. System uptime had a negative association with the weekly questionnaire sum. These results may suggest participants were getting more sleep, despite the weekly score seemingly being associated with an increase in installed apps.

## Discussion

### Principal Results

The results from the SHC 2.0 study over this period did not yield as interesting results as an initial analysis of our SPE study data^30^, likely due to the smaller sample size (N=86 compared to N=315) and reduced number of observations (N=6518 compared to N=25985) in comparison. However, some significant associations were still identified.

While there was a reasonable amount of data collected via the daily questionnaire, some results showed small effect sizes and were occasionally conflicting where there were multiple questions for the same measure, such as for resilience and anxiety. System uptime had a negative association with life satisfaction. Physical activity (app steps) appeared to have a negative association with daily psychological well-being, although this may be related to the small sample size of the study. Daily anxiety results were conflicting. The first measure suggested installed app count had a negative association with daily anxiety; the second measure showed the opposite of this. Other findings, such as the positive associations with app steps, locations visited, floors walked, proximity to campus, were minor. Daily resilience did appear to be associated with proximity to campus and negatively associated with use of the SHC 2.0 app. There also appeared to be a positive association with device uptime, although other results were conflicting. Daily community connectedness appeared to be associated with proximity to campus. System uptime had a positive association with daily depression. There were some significant associations with device digital measures and the daily questionnaire sum, although none were related to physical activity.

For the weekly questionnaire, certain interesting results were found. For instance, device sleep time and install app count had a positive association with weekly life satisfaction. Device sleep time and daily calendar events had a positive association with physical activity. This may indicate participants were getting more sleep or just using their devices less when physically active. The significant associations found with both weekly resilience questions were inconsistent. Floors walked, installed app count, and calendar counts had a negative association with weekly anxiety. Like the daily questionnaire, there were some significant associations between device digital measures and the weekly questionnaire sum, although none were related to physical activity. For three weekly questions (2 - psychological well-being, 5 - depression, and 6 - community connectedness) the mixed linear model fits did not yield any significant associations.

### Limitations

The models have some limitations which likely impacted the results. Missingness did vary on a weekly/daily basis and it likely impacted the results. For instance, there were about 2500 complete case observations from 46 participants daily and 1000-1500 complete case observations from 40-60 participants weekly. The sample size is relatively small compared to the total population of a mid-size university in Canada, although there are a reasonable number of observations for the participants. We were unable to analyze call/text message data from participants due to increasing privacy restrictions on Android/iOS.

### Conclusions

Undergraduate studies (and pandemics) present unique conditions - and opportunities - for surveillance efforts with regards to mental health. This research found that there were some significant associations between phone digital measures and mental health-related self-reports from undergraduates in Ontario, Canada during pandemic conditions. Future opportunities for research include investigations into predictive modelling via machine learning or other means, the comparison of post-pandemic data, and the development of new interventions, such as interactive virtual assistants, or others of ranging complexity, which could potentially incorporate the evidence presented here. While an initial analysis of our related SPE study data^30^ appeared to produce more results as there were considerably more participants and digital measure samples (315 and approximately 25985, respectively) SPE was not designed as a long-term study and has concluded. There remains an opportunity to resume SHC 2.0 and compare data collected during the pandemic to post-pandemic data. Overall, the SHC 2.0 study identified some measures of lifestyle that appear to be associated with measures of mental health in undergraduate students.

## Data Availability

The data for this project is not publicly available as it was an EMA-type study that repeatedly sampled the same participants for potentially sensitive data, such as location.

## Acknowledgements

C.B. thanks Dr. Kevin Shoemaker for initial involvement in Smart Healthy Campus, and Ms. Arlene Fleischhauer for support of that work. C.B. thanks Dr. Lorie Donelle for suggestions to the EMAX software that were incorporated into the SHC 2.0 app. C.B. also thanks Dr. Eva Pila and MacLean Press for contributions to the SHC 2.0 study and app.

## Contributorship

The authors provided have the entire authorship of this article.

## Ethical approval

Ethics approval was provided by Western University’s Health Sciences Research Ethics Board (HSREB) in October 2020 (Project ID 116670).

## Informed consent

Consent was obtained from all research participants prior to the commencement of the research.

## Supplemental material

Supplemental material for this article is available online.

## Abbreviations

CPU: Central Processing Unit
EMA: Ecological Momentary Assessment
EMAX1: Ecological Momentary Assessment eXtensions 1st edition software
EMAX2: Ecological Momentary Assessment eXtensions 2nd edition software
SHC: Smart Healthy Campus
SHCv2: Smart Healthy Campus Version 2 study
SPE: Student Pandemic Experience

